# Changes in stillbirths and child and youth mortality in 2020 and 2021 during the Covid-19 pandemic

**DOI:** 10.1101/2023.09.13.23295484

**Authors:** Enrique Acosta, Lucia Hug, Helena Cruz-Castanheira, David Sharrow, José Henrique Monteiro da Silva, Danzhen You

## Abstract

**Background:** The COVID-19 pandemic’s impact on mortality, especially among the elderly, has been extensively studied. While COVID-19 rarely causes direct mortality in children and youth, the pandemic’s indirect effects might harm these age groups. Yet, its influence on stillbirths and mortality rates in neonates, infants, children, and youth remains poorly understood. This study examines disruptions in such trends across 95 countries in 2020 and 72 in 2021, providing the inaugural comprehensive analysis of COVID-19’s effect on young mortality and stillbirths.

**Methods:** We estimate expected mortality levels in a non-pandemic setting and calculate relative mortality changes (p-scores) by applying generalized linear models to data from civil registers and vital statistics systems (CRSV) and from the Health Management Information System (HMIS). We then use these estimates to analyze, for each age group, the distribution of country-specific mortality changes and the proportion of countries experiencing mortality deficits, no changes, and excess.

**Results:** For most countries and territories, stillbirths and mortality at ages under 25 did not differ from expected levels in 2020 and 2021. However, when focusing on the countries that did show changes, more countries experienced mortality deficits than excess. The exception was stillbirths in both years and mortality among neonates and those aged 10-24 in 2021, where more countries had an excess rather than a deficit. Overall, a quarter of the countries examined experienced increases in stillbirths and young adult mortality (20–24).

**Conclusion:** Despite global disruptions to essential services, stillbirths and youth mortality were as expected in most countries, defying expectations. However, this doesn’t dismiss hypotheses suggesting delayed adverse effects on the youngest that may require more time to be noticeable at the population level. Close and long-term monitoring of health and deaths among children and youth, particularly in low-income and lower-middle-income countries, is required to fully understand the lasting impacts of the COVID-19 pandemic.

**Key messages:**

- This study aims to assess the global impact of the COVID-19 pandemic on stillbirths and child and youth mortality during the years 2020 and 2021.
- We found that despite the pandemic severity and the related socioeconomic disruptions worldwide, most of the 95 countries and territories under analysis experienced no changes in stillbirths and under-25 mortality.
- These findings are important because the impact of the pandemic on the youngest ages remains poorly understood; it contributes essential information for conceiving tailored interventions that can effectively mitigate the adverse consequences of the pandemic on children and youth; and highlight the urgency of strengthen surveillance systems for monitoring health and deaths among children and youth, particularly in low-income and lower-middle-income countries.

## Introduction

In 2020 and 2021, more than 6.5 million COVID-19 deaths were officially reported worldwide. It is well established that the risk of death from COVID-19 increases exponentially with age (1,2), the pre-existence of comorbidities, and social disadvantages (3). It has also been established that official records bias the actual toll of the pandemic due to a lack of testing and the misclassification of causes of death (4,5). A great deal of research aims to assess the overall mortality outcomes of the pandemic (6–9). However, most of these analyses have focused on mortality at old ages, while very little is known about the total impact of the pandemic on pregnancy outcomes and mortality among the youngest age groups.

Deaths caused directly by COVID-19 are rare at young ages (10) and are mainly related to children with severe pre-existing health problems (11,12). According to official reports from countries for which information on direct COVID-19 deaths by age is available, as of the end of 2022, only 0.6% (30,092) of the total number of confirmed COVID-19 deaths were of individuals under age 25 (13). However, children, adolescents, and youth may be indirectly affected by the pandemic, as various pandemic-related disruptions could have adversely impacted their health and well-being. First, one of the earliest consequences of the pandemic was the saturation of healthcare systems in most countries, which resulted in numerous disruptions (14–16). Second, with notable variations from one country to another, most governments adopted several non-pharmaceutical strategies, such as lockdowns, to mitigate the spread of infections. In several cases, these measures drastically disrupted food supply chains, daycare centers and schools, transportation, and many other social institutions and services, leading to the most significant worldwide economic recession in decades (17,18). These and other developments might have negatively affected the physical and mental health of pregnant women, children, and adolescents during the COVID-19 pandemic. It has been hypothesized that the disruption of healthcare systems and decreased access to food during the pandemic exacerbated pre-existing undernutrition levels and other vulnerabilities (19), particularly in low- and middle-income countries (20).

Increases in stillbirth rates have been previously detected during the pandemic in several populations representing diverse socio-economic contexts (21–24). It is likely that these increases were directly or indirectly related to the pandemic. There is evidence that pregnant women were at increased risk of experiencing complications from COVID-19, which, in turn, increased their risk of adverse perinatal outcomes (25). On the other hand, it has been suggested that a deterioration in intrapartum care during the pandemic might have led to an increase in stillbirth risks for pregnant women, irrespective of whether they had been infected with COVID-19 (21,24).

Despite the potential risks to the well-being of unborn infants, children, and youth during the COVID-19 pandemic, analyses of the total impact of the pandemic on mortality at the youngest ages are scarce. This study aims to analyze the changes in stillbirths and mortality among people under 25 during the COVID-19 pandemic in 2020 and 2021. We estimate these changes by measuring variations in stillbirths and child and youth mortality rates relative to the expected levels in a pandemic-free scenario. The difference between the observed and expected all-cause mortality is commonly referred to in the literature as *excess mortality*. As we analyze deficits and excess, we refer to the differences between observed and expected levels as *changes* or *disturbances*.

## Data and Methods

### Data

#### Civil registration and vital statistics systems data

Data consist of annual counts of stillbirths (at 28 weeks or more of gestation) and deaths for seven age groups (i.e., neonates [under 28 days of age], infants [under one year of age], and the age groups 1-4, 5-9, 10-14, 15-19, and 20-24) between 2015 and 2021 obtained from civil registration and vital statistics systems (CRVS). For a robust estimation of expected stillbirths and mortality levels and their corresponding uncertainty, we restricted our analysis to countries with available data for at least three years during 2015-2019 and a population aged under 25 of at least 500,000. Moreover, we excluded data from Armenia and Azerbaijan because the Second Nagorno-Karabakh War heavily affected youth mortality in 2020. Given these criteria, we found CRVS data for 82 countries in 2020 and 57 in 2021.

We retrieved data on live births, stillbirths, and mortality from country-specific CRVS, supplied by countries to UNICEF in response to a data call, and through several databases, including the Human Mortality Database (HMD) (26), the short-term mortality fluctuation data series (STMF) (26), DemoData (27), Eurostat (28), the World Health Organization Mortality Database (WHO-MDB)(29), and the Short-Term Fertility Fluctuations Data Series (STFF) (30). Mortality data are available in death counts; in the cases of Bangladesh, China, India, and South Africa, data were provided either in death rates (*m_x_*) or death probabilities (*q_x_*).

In many cases, we found more than one source for several countries and territories. The criteria for selecting a source were having the most detailed age grouping resolution and the broadest period coverage. When more than one source met these criteria, we prioritized the sources according to the following order: HMD, STMF, country-specific CRVS, DemoData, Eurostat, and WHO-MDB.

Annual population estimates by age group between 2015 and 2021 were obtained from the HMD (26), for the populations for which this information was available, and from the World Population Prospects 2022 (WPP) (31) for the rest. We used data on annual live births for 91 countries in 2020 and 68 in 2021 to account for changes in fertility during the pandemic.

#### Health Management Information System data

We also analyzed monthly Health Management Information System (HMIS) data from 15 low- and lower-middle-income countries in sub-Saharan Africa (12) and Southeast Asia (3). These data include information on stillbirths (15 countries in 2020 and 2021) and deaths of neonates (14 in 2020 and 13 in 2021), and children (9 in 2020 and 8 in 2021). Table S2 in the supplementary materials presents the number of countries with available data by age and income level for 2020 and 2021. Although HMIS data comprise administrative public hospital data — typically with lower coverage than civil register data — we expected to identify signals of mortality change, since underreporting is likely consistent across observed years. Because the available observation period for HMIS data is considerably shorter than for CRVS data (in most cases, from January 2018), we used monthly data for a more robust baseline estimation.

Table 1 presents the number of countries and territories included in the primary analyses with CRVS and HMIS data on births, stillbirths, and deaths by age group for 2020 and 2021. Countries were categorized by income level according to the most recent World Bank income classification (32). Tables S1-S4 in the supplementary materials present detailed information on CRVS and HMIS data availability and sources by demographic measure and country.

**Table 1.**
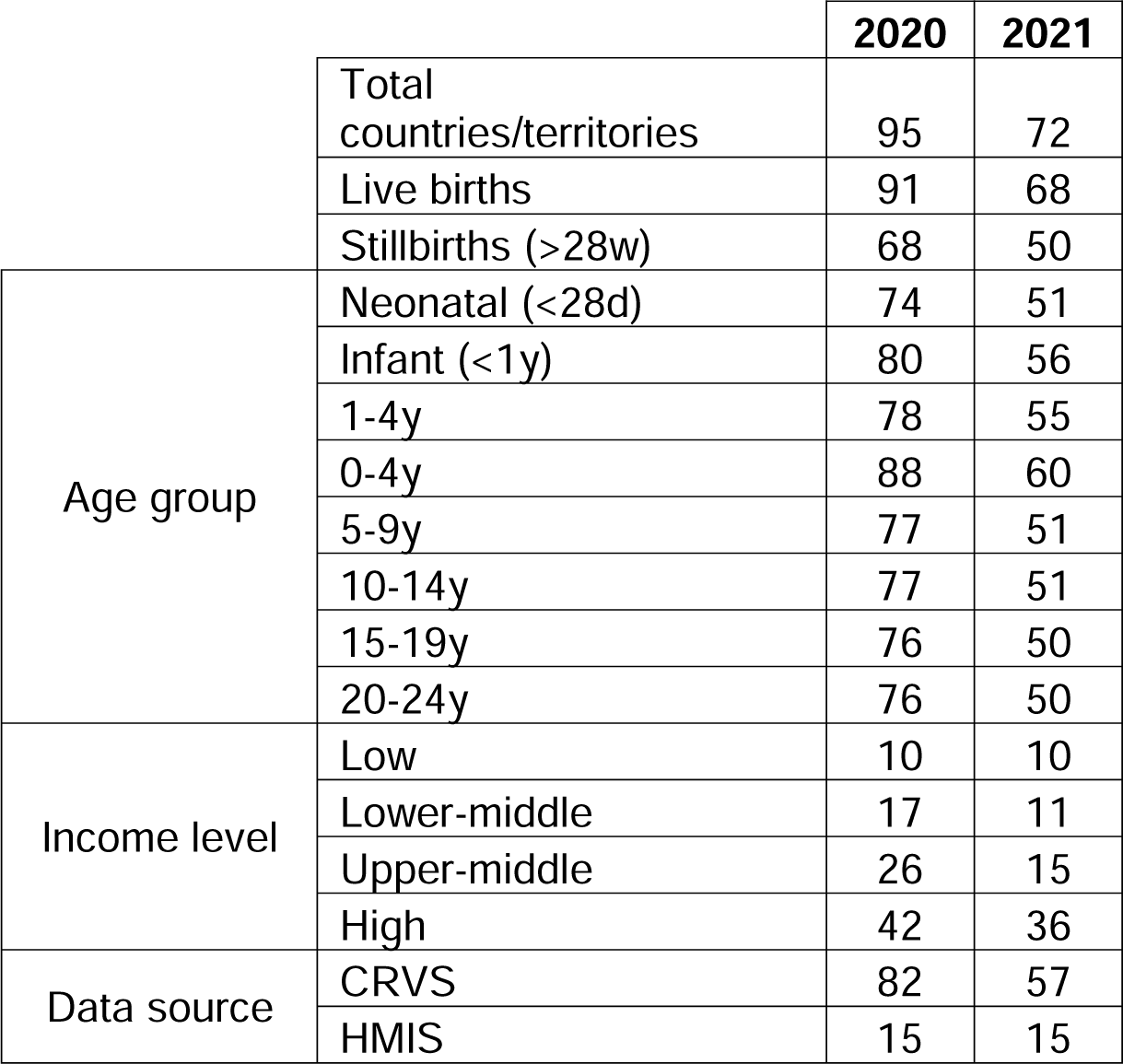
Summary of the number of countries and territories included in the analysis with data on live births, stillbirths, and mortality under 25 by age, income level, and data source in 2020 and 2021.

The world map in Figure 1 presents the geographical distribution of the countries for which CRVS or HMIS data are available for 2020 and 2021. The maps in the supplementary materials (Figures S1-S4) give additional information on data availability by demographic measure, income level, and year.

**Figure 1.**
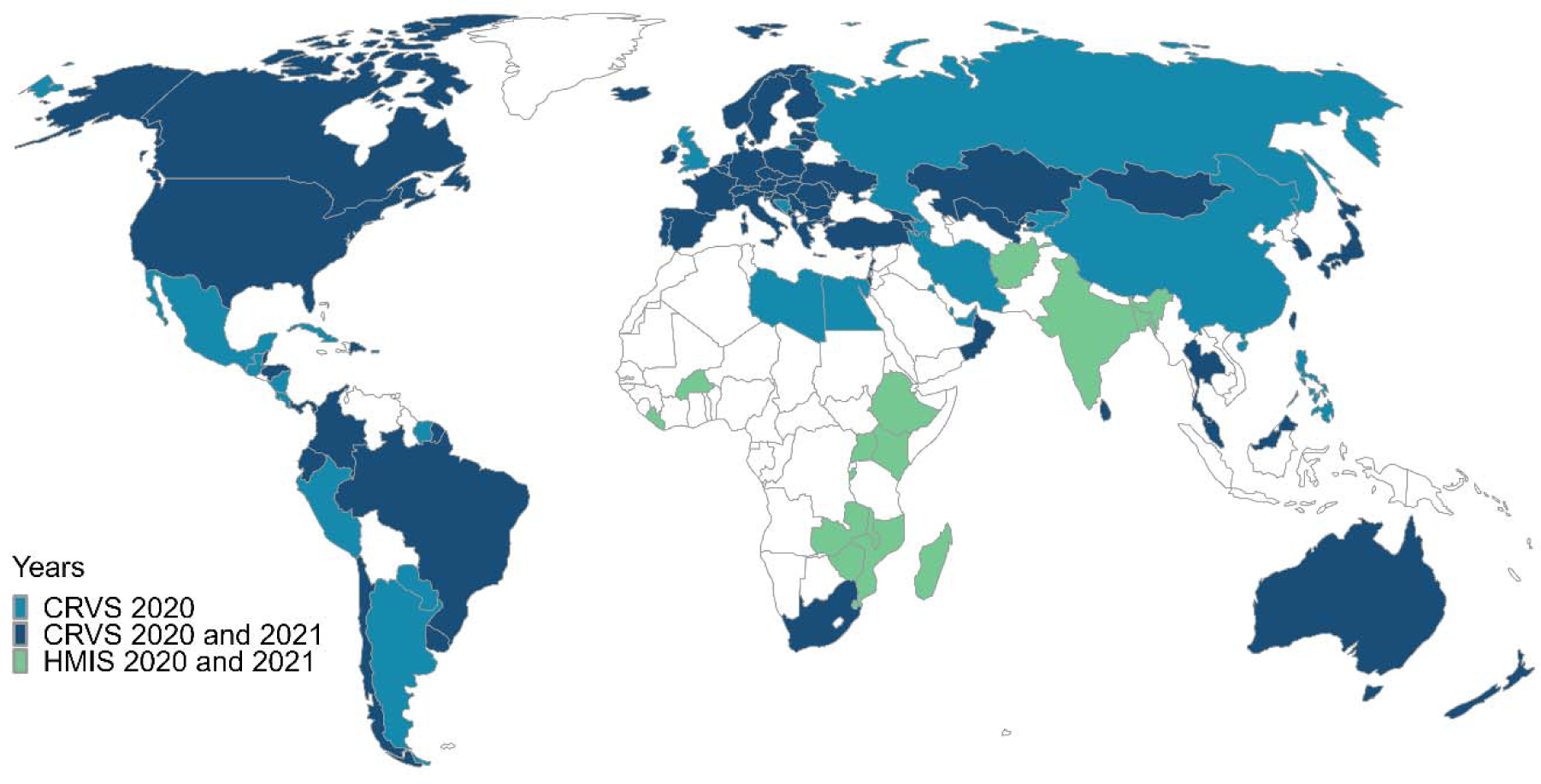
Countries and territories with available CRVS and HMIS data on stillbirths or all-cause mortality at any 5-year age group under 25 for 2020 and 2021. Besides HMIS data, Bangladesh also had CRVS data on infant and child mortality in 2020 and India on infant mortality in 2020.

## Methods

We define mortality change as the difference between the observed all-cause mortality and the expected mortality “in the absence of the pandemic” (also denoted as baseline mortality) during 2020 and 2021. The method we used for CRVS data accounts for secular changes in mortality, population size, and age structure over time. We obtained the baseline mortality by fitting a country- and age-specific generalized linear model to annual death counts between 2015 and 2019. The model uses a quasi-Poisson likelihood to account for overdispersion. We used this model to predict the expected deaths for 2020 and 2021. The model is defined as

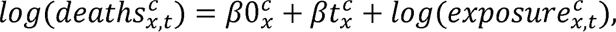

where 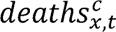 and 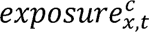 indicate the death counts and population at risk for each age group *x* and country *c*, in year *t*. The term 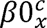 accounts for the intercept and 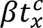 for secular changes in mortality (as an exponential trend) by age and country. We estimated 95% upper and lower prediction intervals by bootstrapping, using 2,000 simulations. Examples of the model fitting are available in the supplementary materials (Figures S4-S5).

We obtained a monthly baseline for countries with HMIS data by fitting a generalized additive model to include a cyclic spline component accounting for within-annual seasonality. Health Management Information System monthly baseline estimates were then aggregated to compute annual relative changes and uncertainty levels. The supplementary materials present additional details on the model for monthly baseline estimation. We present analyses and findings from CRVS and HMIS data separately due to the substantial differences in data characteristics and methods we applied to each case.

After obtaining baseline mortality estimates, we computed the *p-score* index (33) to measure mortality changes in 2020 and 2021. P-scores indicate the relative change in observed mortality compared to the baseline, expressed in percentage. The p-score index for each country *c*, age *x*, and year *t* is calculated as

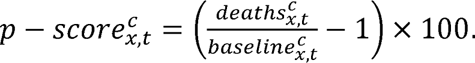

Using the p-score index to measure mortality changes has two main advantages. First, it has a straightforward interpretation as the percentage change in mortality relative to the expected value in the absence of the pandemic. Second, it allows us to compare mortality changes across countries, regardless of differences in mortality levels and population sizes.

We also estimated age-specific *overall p-scores* as a summary measure of excess mortality for all of the populations examined together. Overall p-scores were obtained by adding all available death counts and population exposures together in the numerator and denominator. Given that it is necessary to have complete data since 2015 to compute overall p-scores, we included 95 and 72 countries and territories (without restrictions regarding population size) for 2020 and 2021, respectively.

## Results

Figure 2 presents two summary plots of the age-specific p-score estimates for the **82** countries with CRVS data included in the analysis. Figure 2A plots the distribution of all country-specific p-score estimates by age group for 2020 and 2021, indicating whether they resulted in deficits, no changes, or excess. Although p-score values vary widely across countries and age groups (between −58% and 123%), the p-score interquartile ranges (indicated with horizontal black bars) are spread between −28% and 27% in all observed age groups. Figures S7 and S8 (see supplementary materials) depict the p-score estimates for each age group and country with CRVS data.

**Figure 2.**
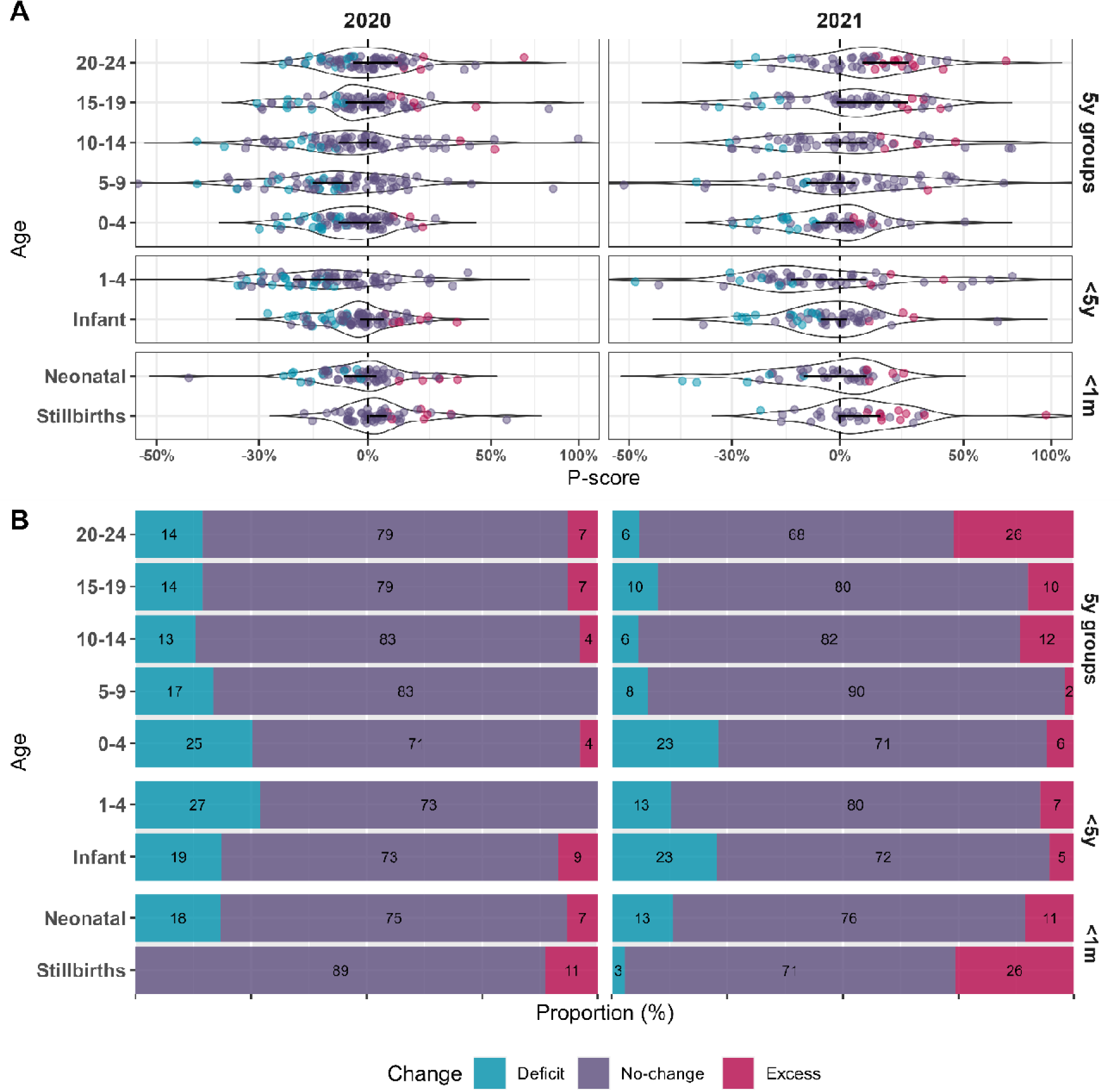
Summary of p-score estimates in 2020 and 2021 by age group among countries and territories with CRVS data. Panel (A) depicts the distribution of country-specific p-score values by age group. Black dots and horizontal black bars indicate the population-weighted median and 25th and 75th percentiles of the p-score distributions. Panel (B) plots the proportion (in percentage) of estimates resulting in deficits, non-significant changes, and excess mortality.

Figure 2B plots the proportion of countries and territories with CRVS data showing mortality deficits, non-significant changes, and excess by age group and year. According to this plot, across all observed age groups, most countries (68% to 90%, as indicated by the purple bars) experienced mortality as expected in 2020 and 2021.

However, besides the overall pattern of non-significant changes in mortality, as presented in Figure 2, it is enlightening to focus on the composition of excess (indicated in red) and deficits (in blue) among the countries that did experience changes in mortality. Stillbirths were the only group in which, among the countries showing changes, more countries experienced an excess rather than a deficit in both years. In 2020, across all age groups, with the exception of stillbirths, deficit deaths were considerably more prevalent than excess among countries experiencing mortality changes. The landscape is different in 2021, where, besides stillbirths, excess mortality was also more frequent than deficits for ages 10-14, 15-19, and 20-24, again among the countries experiencing changes. Remarkably, overall in 2021, one in every four countries witnessed increases in stillbirths and one in every four saw an increase in deaths at ages 20-24 — though these two observations are not necessarily true for a single country simultaneously. Figures S9 and S10 (see supplementary materials) present the p-score estimates distribution and proportion by income level. These results suggest a similar pattern of stillbirths and mortality changes across the income levels under observation: the considerably predominant tendency is toward countries experiencing no changes in mortality, with a weak tendency toward mortality deficits in 2020 and increases in excess for some ages in 2021. Regardless of income level, more countries experienced excess than deficits in stillbirths and deaths at ages 20-24 in 2021.

Figure 3 presents summary p-score estimates for the 15 countries with HMIS data. The pattern of these results is similar to the one observed among countries with CRVS data in Figure 2 in at least two aspects. First, in most countries, stillbirths and mortality rates were as expected. Second, a greater number exhibited excess rather than deficits in the countries where stillbirth changes were observed in both years.

**Figure 3.**
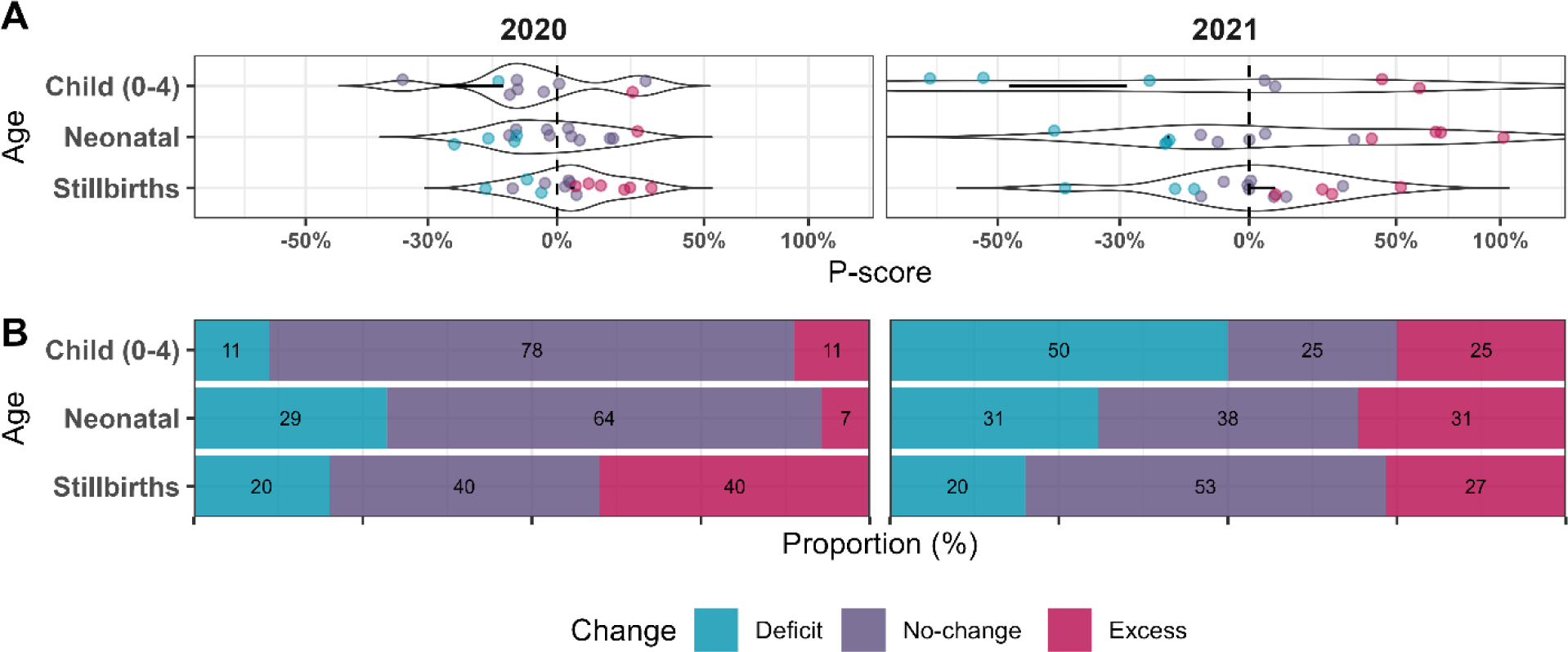
Summary of p-score estimates for 2020 and 2021 for stillbirths and neonatal and child mortality among countries and territories with HMIS data. Panel (A) depicts the distribution of country-specific p-score values by age group. Black dots and horizontal black bars indicate the population-weighted median and 25th and 75th percentiles of the p-score distributions. Panel (B) plots the proportion (in percentage) of estimates resulting in deficits, non-significant changes, and excess mortality.

Focusing on the analysis of mortality changes in all observed countries and territories as one unique population, Figure 4 presents the age-specific overall p-scores for countries with complete CRVS data between 2015 and 2020 (left panel) and between 2015 and 2021 (right panel). The fitting of the overall baselines is presented in Figure S11 in the supplementary materials. The age pattern of the overall p-score estimates, presented in Figure 4, is consistent with the country-specific estimates in Figure 2. Regardless of whether mortality change is computed separately by country or for all countries with sufficient data as a whole, our findings consistently suggest that unborn children and young adults aged 20-24 are the most vulnerable age groups under analysis, showing slight signals of deterioration during the pandemic.

**Figure 4.**
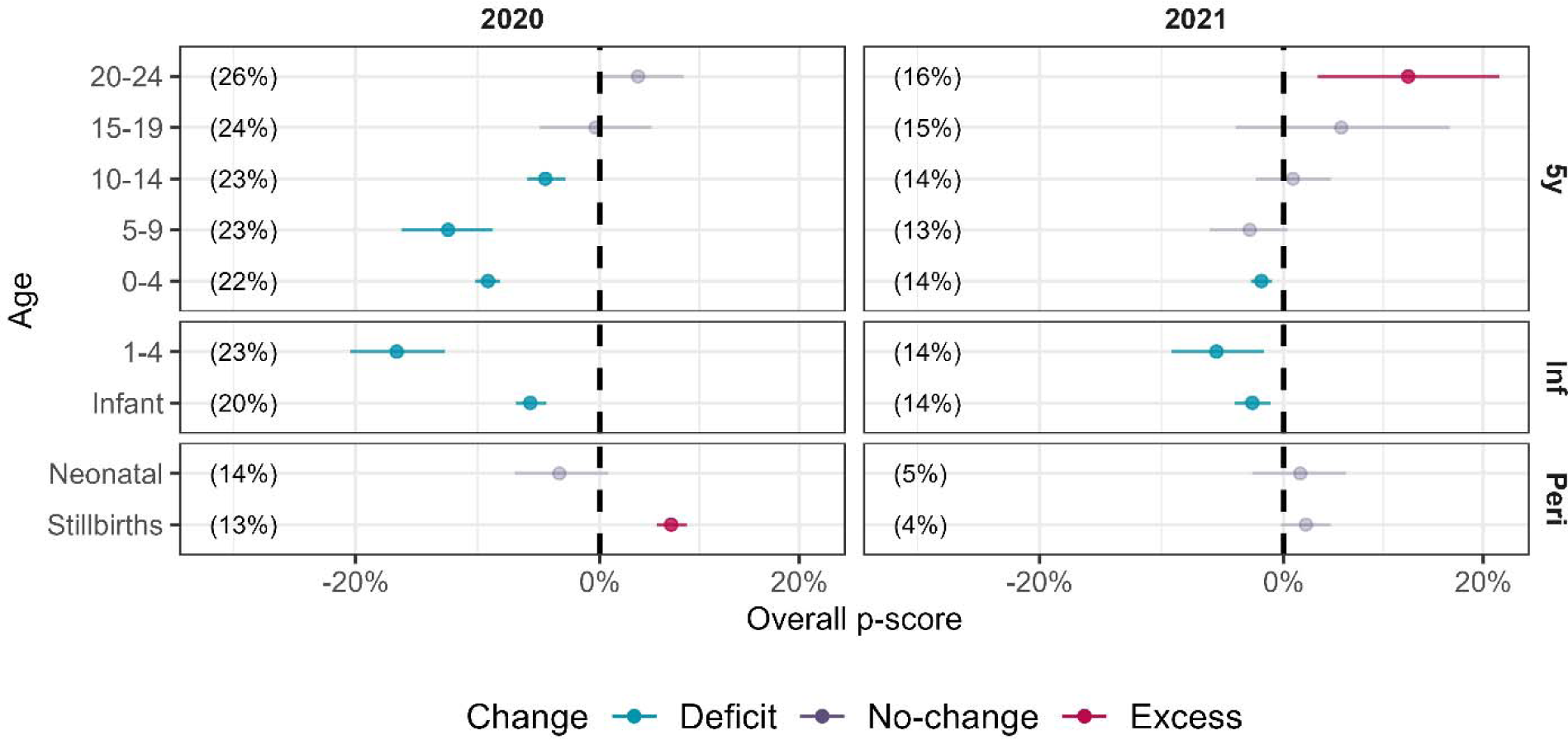
Overall p-scores in 2020 and 2021 by age. Vertical dashed lines indicate no change in mortality relative to the expected value (i.e., p-score of 0%). Numbers in parentheses indicate the percentage of the total world population considered in the analysis within each age group.

The percentages of the world population covered in this analysis, indicated in parentheses by age group and year, range between 13% and 26% in 2020 and 4% and 16% in 2021. It is essential to once again underline that the representativeness of the population under analysis is directly proportional to the income level. Figure S12 (see supplementary materials) presents the overall p-score estimates by income level. Whereas 94% of the population in high-income countries is included in these analyses for 2020 and 78% for 2021, these proportions decrease steadily in the case of upper-middle (38% and 20%) and lower-middle (8% and 2%) income countries. As mentioned in the data section, we could not include any low-income country in the computation of overall p-scores because of a lack of CRVS data.

## Discussion

We analyzed relative changes in stillbirths and child and youth mortality in 95 countries in 2020 and 72 in 2021. Our findings suggest that, despite the severe health crisis and socioeconomic disruptions experienced during the COVID-19 pandemic, most countries and territories analyzed had the expected stillbirth and mortality levels across all observed age groups during both years. Nevertheless, in the case of countries where changes are observed, there are noticeable differences between both years. While, in 2020, mortality reductions were more frequent than increases in all age groups, with the exception of stillbirths, in 2021, increases were more prevalent than reductions among stillbirths and mortality among those aged 10-24 years. The age pattern of mortality changes and the variations from 2020 to 2021 are highly consistent through the different analyses we perform.

Regarding income levels, we found a similar pattern in the distribution and proportions of relative changes in stillbirths and deaths across lower-middle-, upper-middle-, and high-income countries (see Figure 3 and Figures S8-S9 in the supplementary materials).

Considering the ample evidence of minimal direct adverse effects caused by the COVID-19 disease at young ages, it is highly likely that both estimated excess and deficits were indirectly influenced by the pandemic. In other words, mobility restrictions, isolation, the closure of schools, other non-pharmaceutical strategies adopted by governments, and disruptions in healthcare and food supply systems might have driven the observed stillbirths and mortality perturbations described in this study. Although the mechanisms modulating the observed disturbances in mortality are beyond the scope of this paper, we can hypothesize about them. Our finding regarding stillbirth increases in 11% and 26% of the observed countries in 2020 and 2021 could be related to pandemic-related anxiety (34), nutritional deficiencies, reduced access to or delays in antenatal care, and decreases in preterm births (21–24). However, the mechanisms driving excess stillbirths during the pandemic remain quite speculative, as several studies have reported conflicting evidence, and further analyses are needed (24,35).

The finding that a higher number of countries experienced excess stillbirths than deficits, coupled with a greater prevalence of infant mortality deficits than excess, suggests a potential link through *harvesting effect* mechanisms. Insufficient medical attention and increased anxiety during pregnancy might lead to stillbirth increases (24), disproportionately affecting the frailest fetuses in gestation. This selection pressure might have resulted in a more robust composition of live births with lower mortality risks than in previous years. Nevertheless, although this mechanism seems consistent with our findings that indicate that more countries experienced excess than deficits in stillbirths and infant mortality in 2020, it can neither explain the predominance of mortality deficits we found among children aged 1-9 in both years nor the increase in the proportion of countries with excess neonatal deaths in 2021.

The increases in mortality for adolescents (10–19) in one-tenth of the observed countries in 2021 and in one-fourth for young adults (20–24) could be associated with pandemic-related psychosocial stressors that disproportionately affected these ages. Many children and youth at developmentally sensitive life stages suffered prolonged social isolation — in many cases trapped in dysfunctional family settings —, loss of social support and safety nets, and increased economic instability, among other stressors (36,37). These risks were considerably higher for specific populations, such as sexual and gender minorities, and youth suffering from substance abuse and other pre-existing psychological disorders (38–40). In addition to these indirect mechanisms affecting young ages, it is also plausible that a fraction of the excess mortality we found in 2021 for ages 20-24 was directly caused by COVID-19. There is evidence that the new SARS-CoV-2 virus variants that emerged in 2021 had increased risks of developing severe outcomes at young ages (41,42).

### Sensitivity analyses

We performed three robustness checks to evaluate how sensitive our estimates are to the inclusion of small population countries and territories and the time resolution of data, as well as to test how exceptional our findings are to the pandemic years.

First, we tested the impact of including countries and territories with relatively small youth populations (under 500,000 children and youth under 25). Table S4 (see supplementary materials) presents the available information by age group and year for this robustness check. Figure S14 compares weighted age-specific p-scores average values and ranges for 2020 and 2021, depending on the inclusion or exclusion of countries with small populations. According to this analysis, the weighted p-scores averages are unaffected, but, as expected, the range of values increases when including small populations. Second, we also compared our annual-based estimates to those obtained using weekly data, where possible, to test how sensitive our results are to differences in time resolution modeling. The results of this comparison, presented in Figure S15 in the supplementary materials, suggest that our findings are robust and not dependent on the time resolution of data. Finally, we estimated mortality changes for the years 2017, 2018, and 2019 in 75 countries, using the preceding five years in each case to estimate the baseline mortality. Figures S16-S18 in the supplementary materials present mortality change estimates for 2017, 2018, and 2019. These results suggest that the perturbations observed in 2020 and 2021 are exceptional to the pandemic context and did not result from random fluctuations in mortality that could occur in any given year.

### Limitations

We acknowledge several limitations. First, due to data availability, our investigation suffers from a substantial socio-economic bias towards high- and upper-middle-income countries. The efficient and adequate collection and publication of register and vital statistics data require considerable human and physical resources. Unfortunately, most lower-middle- and low-income countries cannot release rapid mortality updates that are sufficiently reliable to analyze nationally representative short-term mortality fluctuations. However, our analyses of HMIS data from low- and lower-middle-income countries suggest strong similarities to the CRSV data findings in the magnitude and direction of relative changes in stillbirths and child mortality: in both sets of data, most countries showed no changes in stillbirths or mortality. Among the countries with observed variations from expected levels, excess stillbirths were more frequent than deficits, and child mortality deficits occurred more frequently than excess.

A second limitation pertains to data quality, particularly with regard to delays and the under-registration of vital events. Registration delays in 2020 and 2021 might have biased mortality change estimates downwards. However, we consider that enough time has passed to allow for adjustments to the registration of deaths that occurred in 2020, where estimates resulted in even larger deficits than those obtained for 2021. Although further inclusion of delayed events registration might increase the values in all-cause mortality and mortality change estimates, we do not expect those adjustments to alter our findings substantially. Regarding the under-registration of vital events, as the p-score index measures mortality changes in relative terms, constant levels of under-registration do not lead to bias in our estimates. We have not found evidence suggesting that vital statistics coverage was substantially modified during 2020 or 2021, although this is possible for several low- and middle-income countries. Despite the possibility of delayed registrations, we do not expect that our findings result from data artifacts because high- and upper-middle-income countries — with high-quality data — are over-represented in our analysis.

Identifying the mechanisms that modulated the observed changes in mortality requires different data and methods from the ones used in this study. For instance, further analyses on changes in the composition of mortality by cause of death would be required to better understand the mechanisms affecting stillbirths and deaths among children and youths.

## Conclusion

We found no widespread significant changes in stillbirths and mortality among the ages and countries under observation. However, among the countries that experienced significant changes in mortality in 2020, all groups but stillbirths showed lower-than-expected mortality. In 2021, together with stillbirths, neonates and those aged 10-24 also saw a prevalence of excess rather than deficits. In particular, the increase in the proportion of countries with higher-than-expected mortality in 2021 for stillbirths and ages 20-24 (26% in both cases) is noteworthy.

These findings are surprising given the considerable disruption of food supply and healthcare systems during the pandemic. Nevertheless, our findings do not invalidate the hypotheses that predict a detrimental impact of the pandemic on the health of the youngest segments of the population in the mid or long term. These disruptions may take more time to have a noticeable negative impact on mortality at the youngest ages. Close and long-term monitoring of health and deaths among children and youth, particularly in low-income and lower-middle-income countries, is required to fully understand the lasting impacts of the COVID-19 pandemic. This monitoring would require faster and better vital statistics systems, particularly deficient in low- and middle-income countries. Finally, further analyses with additional data on causes of death are needed to fully understand the mechanisms behind the changes in mortality we found in this study. This information will be essential for assessing the effectiveness of governmental responses and strategies intended to mitigate the burden of the pandemic.

## Data Availability

All underlying input data from civil registration systems are openly available databases. All the software code for reproducing the analyses are available in OSF, at https://dx.doi.org/10.17605/OSF.IO/FNUEY

## Authors’ Contributions

EA contributed to conceptualization, data collection and curation, methodology, formal analysis, visualization, and review and editing – original draft. LH, HC, DS, and DY contributed to conceptualization, data collection and curation, and writing – review & editing. JM contributed to data curation.

## Acknowledgments

An earlier version of this article was presented at the 2022 meeting of the Asociación Latinoamericana de Población (ALAP) and the 2023 meeting of the Population Association of America (PAA). We thank the following individuals for valuable interactions during the preparation of this manuscript: Patrick Gerland, Jon Wakefield, Bruno Masquelier, Li Liu, Kenneth Hill, Michel Guillot, Leontine Alkema, and José Manuel Aburto.

