## Supplementary Materials for "Changes in stillbirths and child and youth mortality in 2020 and 2021 during the Covid-19 pandemic"

### DESCRIPTION OF DATA AND SOURCES

#### Summary of data

Table S1 presents the number of populations with available CRVS data on births and all-cause mortality by age group for 2020 and 2021 (columns 2 and 3) and the number of populations with at least 500 thousand inhabitants aged under 25 years, which were included in the primary analyses of this study (columns 4 and 5). Table S2 presents the number of countries with HMIS data included in this study.

**Table S1.** Countries and territories with available annual CRVS data on stillbirths and all-cause mortality by age and income level for the years 2020 and 2021

| | | Available data | | Above population threshold (500K) | |
|---|---|---|---|---|---|
| | | 2020 | 2021 | 2020 | 2021 |
| **Total countries** | | 111 | 71 | 82 | 55 |
| **Age at death** | Stillbirths (>28w) | 67 | 37 | 56 | 31 |
| | Neonatal (<28d) | 70 | 37 | 59 | 32 |
| | Infant (<1y) | 106 | 56 | 80 | 46 |
| | 1-4y | 104 | 55 | 78 | 44 |
| | 0-4y | 106 | 66 | 79 | 50 |
| | 5-9y | 106 | 66 | 77 | 50 |
| | 10-14y | 105 | 65 | 77 | 50 |
| | 15-19y | 103 | 64 | 76 | 49 |
| | 20-24y | 103 | 64 | 76 | 49 |
| **Income level** | Lower-middle | 13 | 5 | 12 | 5 |
| | Upper-middle | 35 | 20 | 28 | 16 |
| | High | 63 | 46 | 42 | 34 |

**Table S2.** Countries with available HMIS monthly data on stillbirths and neonatal and child mortality by income level for the years 2020 and 2021

| | | 2020 | 2021 |
|---|---|---|---|
| **Total countries** | | 15 | 15 |
| **Age at death** | Stillbirths (>28w) | 15 | 15 |
| | Neonatal (<28d) | 14 | 13 |
| | 0-4y | 9 | 8 |
| **Income level** | Low | 10 | 10 |
| | Lower-middle | 5 | 5 |

### Data coverage

Figure S1-S3 presents world maps indicating the countries and territories with available CRVS or HMIS information on live births, infant deaths, and the income levels.

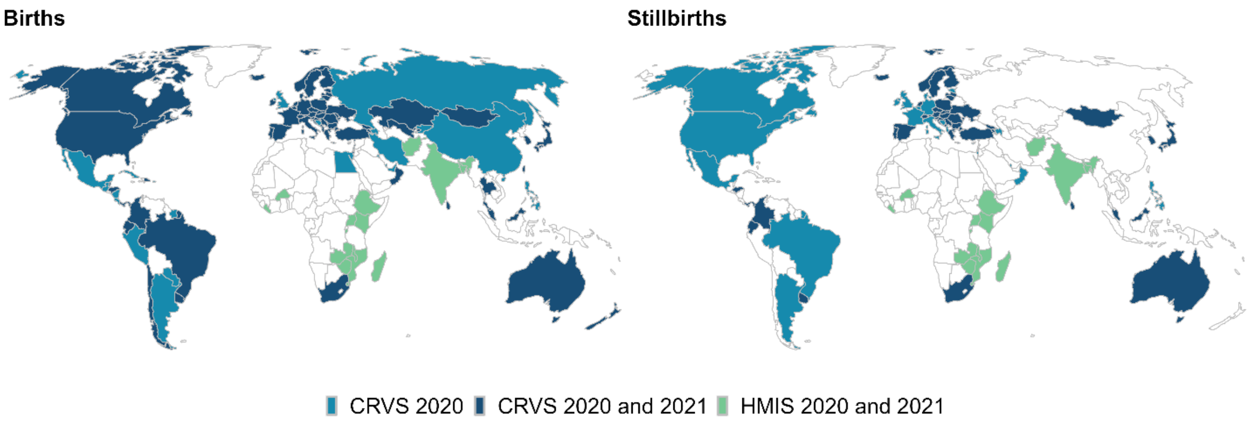

**Figure S1.** Countries and territories with data on births and infant deaths in 2020 and 2021

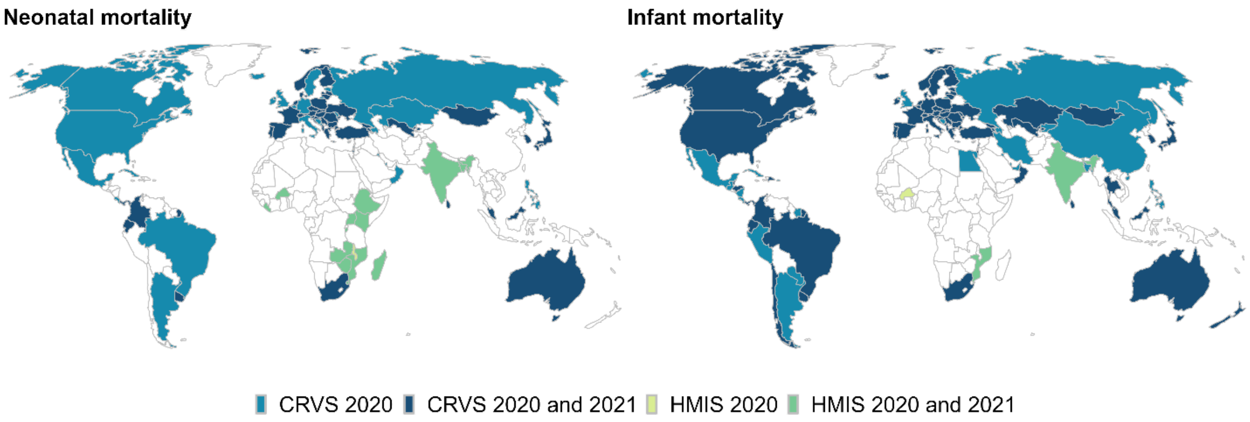

**Figure S2.** Countries and territories with data on stillbirths and neonatal deaths in 2020 and 2021

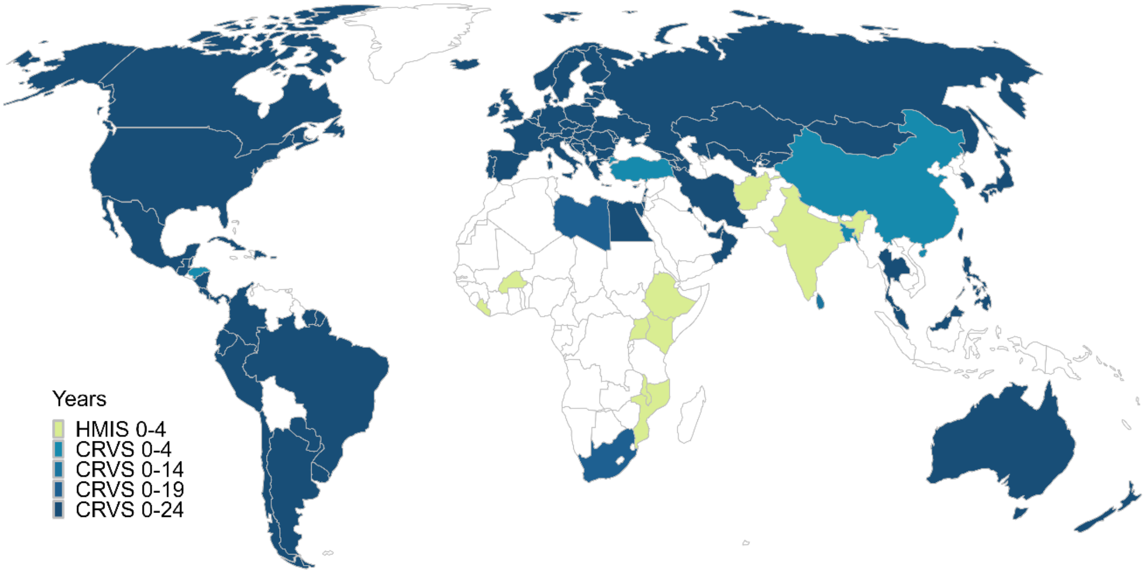

**Figure S3.** Countries and territories with data on mortality for ages 0-24 in 5-year age groups in 2020 and 2021

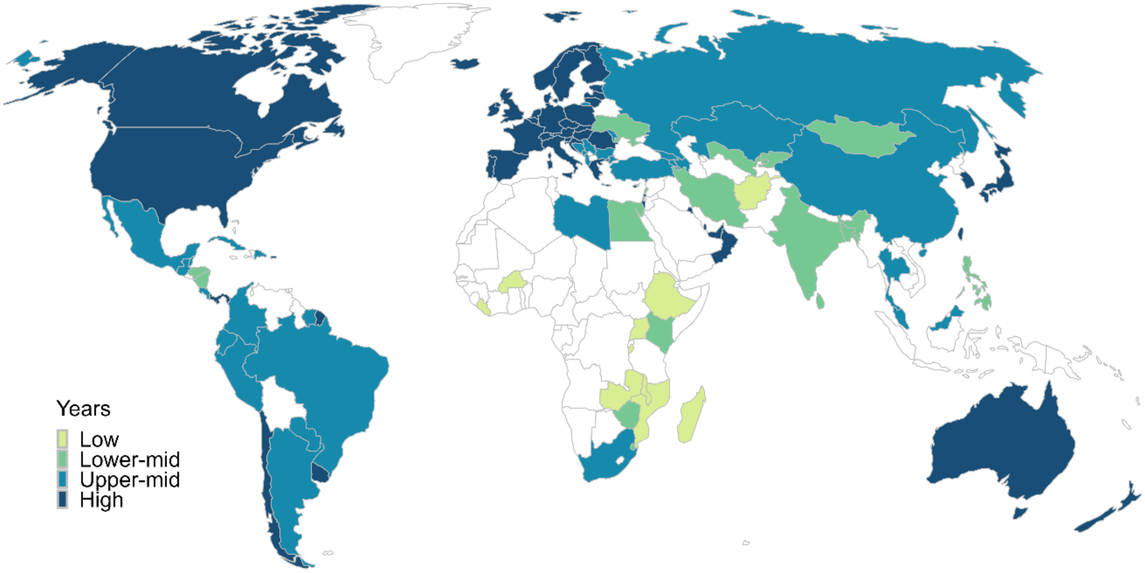

**Figure S4.** Countries and territories with data by income level

### METHODS

#### Model for monthly baseline mortality estimation

We used monthly HMIS data to obtain excess mortality and stillbirths to include more observations. The method we used for modeling monthly data accounts for three components: 1) secular changes in mortality, 2) within-year seasonality, and 3) changes in population size and age structure over time. We obtained the baseline mortality by fitting a country- and age-specific Generalized Additive Model (GAM) to monthly death counts, using a quasi-Poisson distribution to account for overdispersion. The model is defined as

$$log(deaths^{c}_{x,\,t}) = \beta_0 + \beta_1 t + \beta_2 m + log(exposure^{c}_{x,t}),$$

where $deaths^{c}_{x,t}$ and $exposure^{c}_{x,t}$ indicate, respectively, the death counts and population at risk for each age group *x* and country *c*, in year *t*. The term $\beta_0$ accounts for the intercept, $\beta_1 t$, for secular changes in mortality (as a linear trend in the logarithmic scale), and $\beta_2 m$ for within-year seasonality.

#### Examples of annual baseline mortality fitting

We present a few examples of age-specific baseline mortality fitting in four countries and all age groups in Figure S4. Excess mortality is the difference between the expected mortality in the absence of the pandemic (black line) and the observed mortality (red dots) in 2020 and 2021. P-score indexes are estimated as the relative differences and expressed in percentage values to facilitate interpretation and comparisons.

*Stillbirths and neonatal deaths*

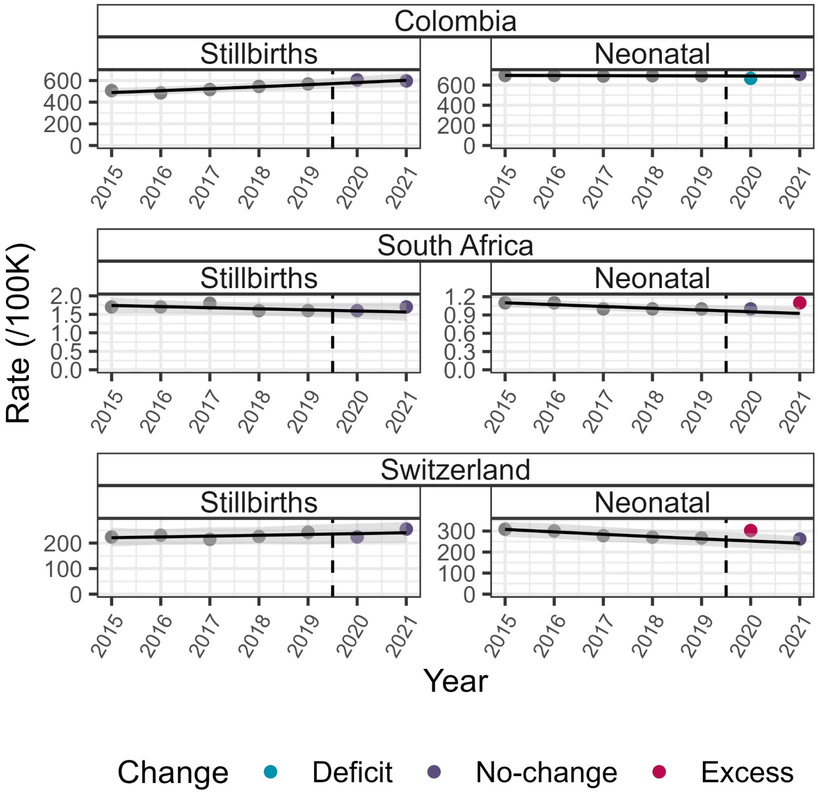

**Figure S5. Baseline mortality fitting by age group for all sexes combined in Colombia, South Africa, and Switzerland, using a Generalized Linear Model with overdispersion.** Black dots indicate observed annual death counts between 2015 and 2019, and colored dots indicate death counts in 2020 and 2021, indicating negative (in blue), positive (in red), or no (in purple) excess mortality. The black line depicts the fitted baseline mortality, and the gray band its bootstrapped 95% prediction interval.

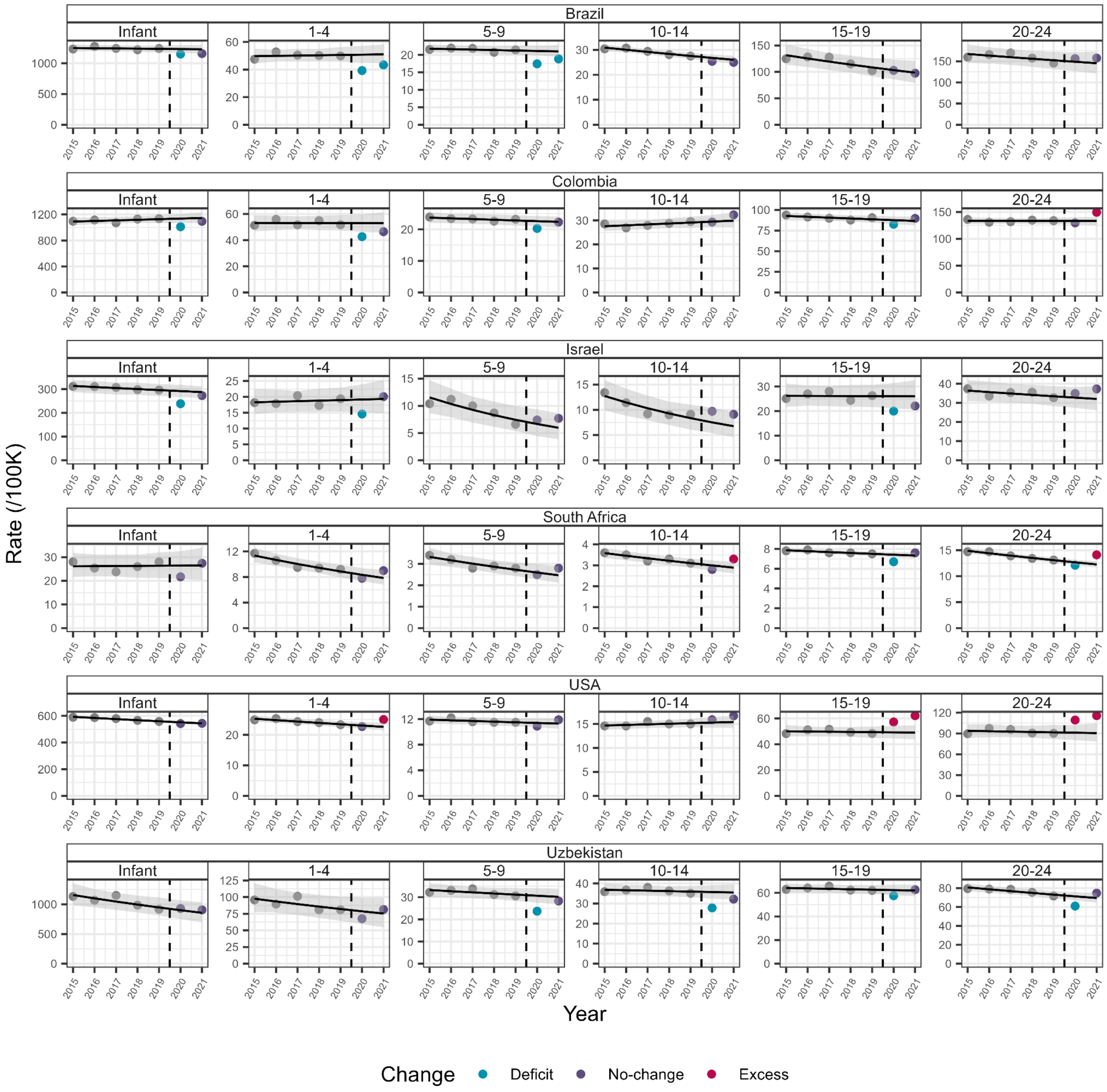

**Figure S6. Baseline mortality fitting by age group for all sexes combined in Brazil, Colombia, Israel, South Africa, USA, and Uzbekistan.** The baseline mortality is obtained by fitting a Generalized Linear Model with overdispersion on observed annual death counts between 2015 and 2019 (black dots). The black line depicts the fitted baseline mortality, and the gray band its bootstrapped 95% prediction interval. Observed annual death counts for 2020 and 2021 are colored, indicating positive (in red), negative (in blue), or no (in purple) excess mortality.

### ESTIMATES

#### P-score estimates by age and country for CRVS data

Figure S5 presents the country-specific p-score estimates for excess stillbirths and deaths by age and income level.

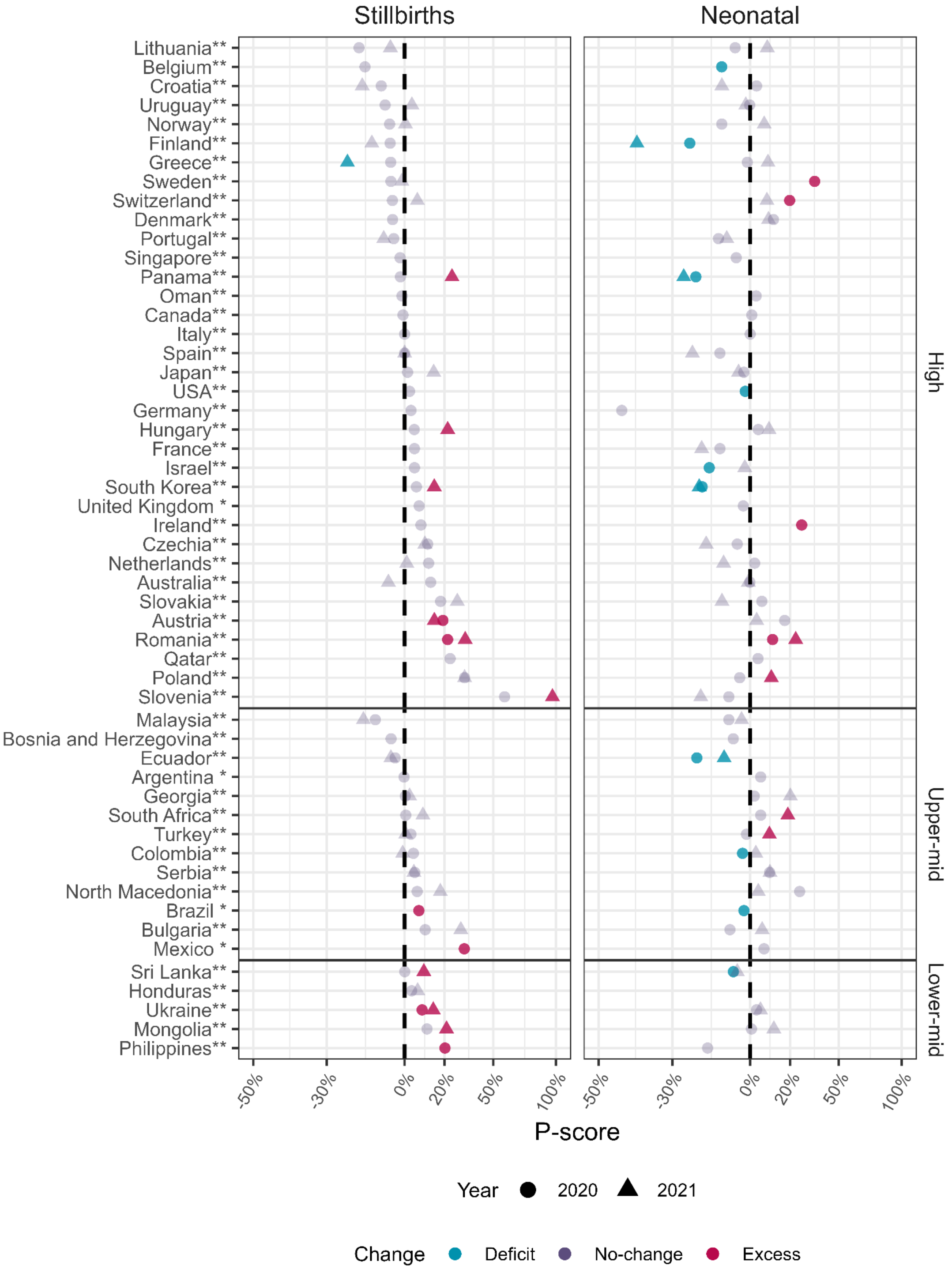

**Figure S7. Age group-specific p-score estimates by country and income level.** Vertical dashed lines indicate no change in mortality relative to the expected value (i.e., P-score = 0%). Points in blue and red indicate negative and positive changes in mortality that are statistically different from 0% (ρ < 0.05), and gray points indicate changes that are not statistically different from 0% (ρ ≥ 0.05).
(*) countries with available data on live births for 2020.
(**) countries with available data on live births for 2020 and 2021.

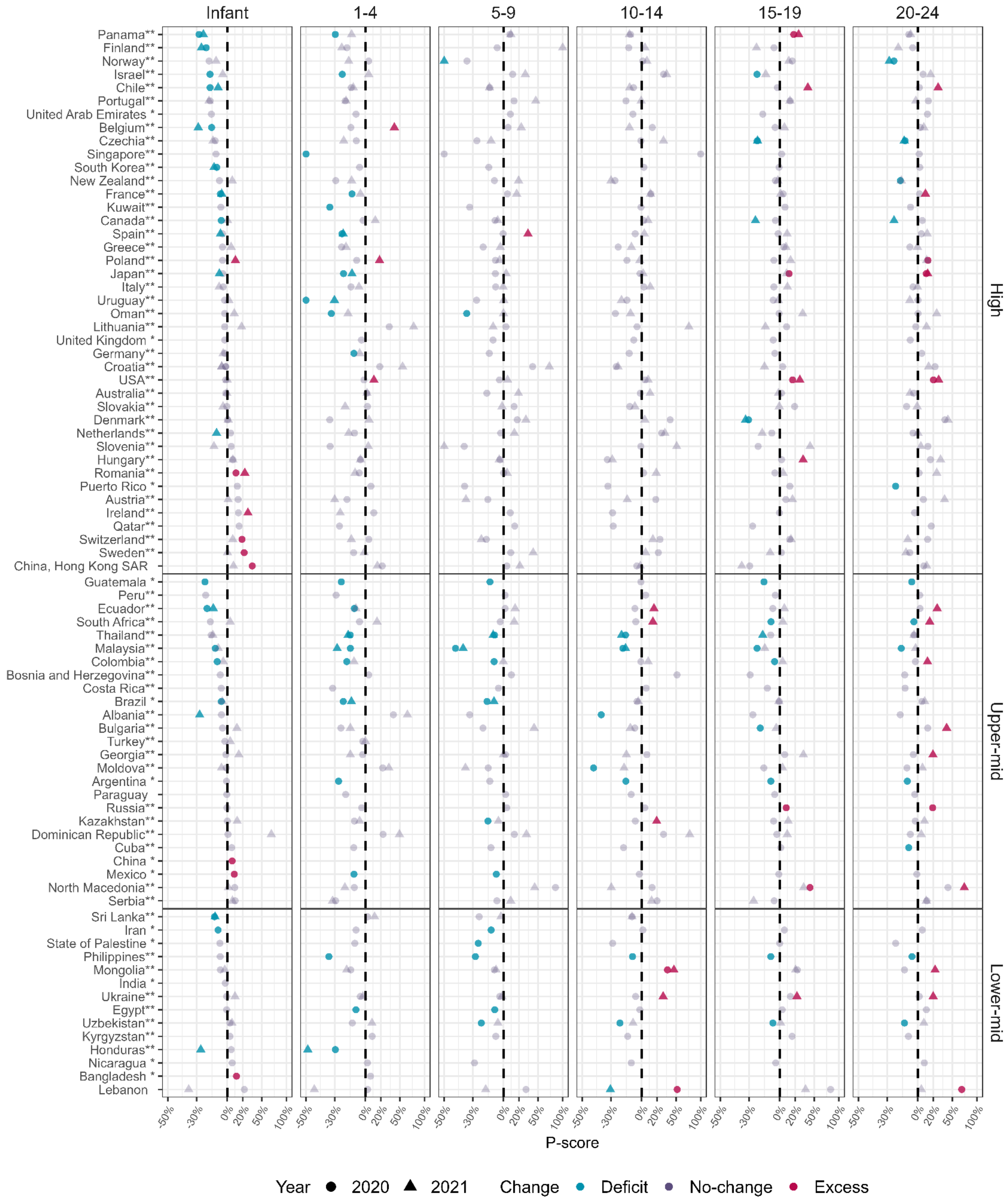

**Figure S8. Age group-specific p-score estimates by country and income level.** Vertical dashed lines indicate no change in mortality relative to the expected value (i.e., P-score = 0%). Points in blue and red indicate negative (deficit) and positive (excess) changes in mortality, respectively, that are statistically different from 0% (ρ < 0.05), and gray points indicate changes that are not statistically different from 0% (ρ ≥ 0.05).
(*) countries with available data on live births for 2020.
(**) countries with available data on live births for 2020 and 2021.

### Distribution of age- and country-specific p-scores by income level

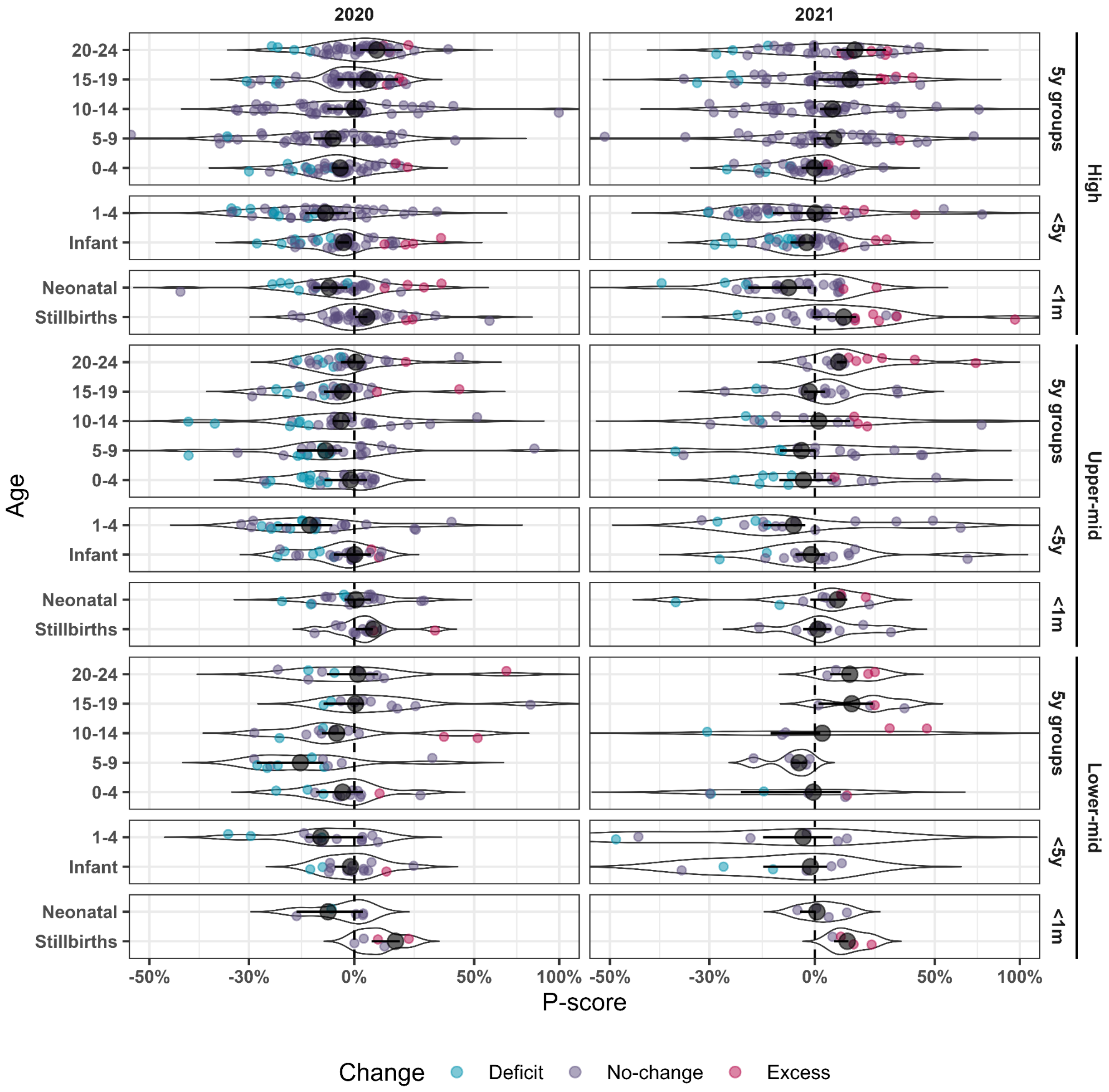

**Figure S9. Distributions of p-score estimates in 2020 and 2021 by age group and income level.** Empty black circles indicate weighted means, full black circles population-weighted medians, and horizontal black bars indicate the 25th and 75th percentiles of the country-specific p-scores distribution.

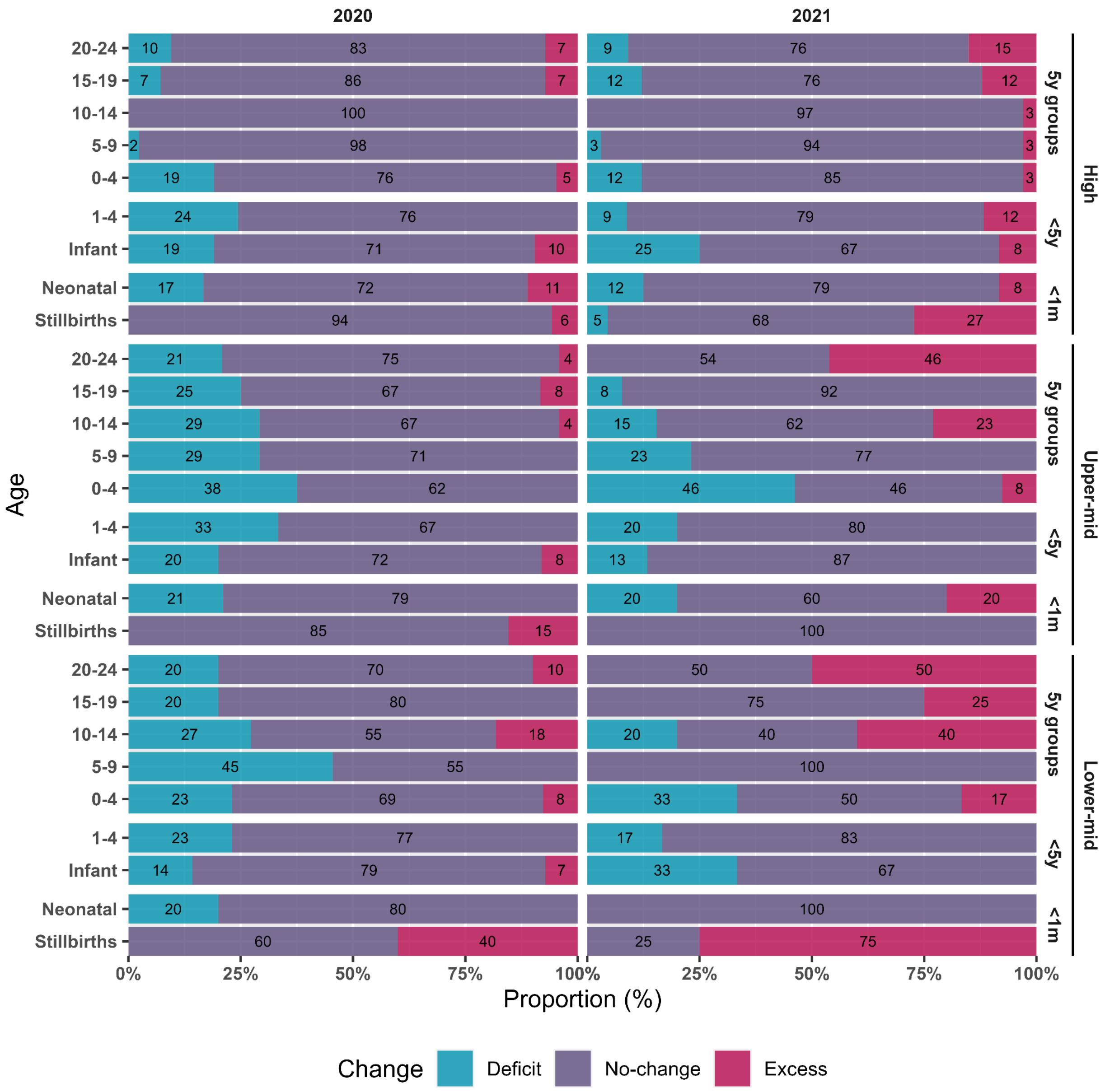

**Figure S10. Proportions of populations according to p-score estimates in 2020 and 2021 by age group and income level.**

### Overall p-scores estimation

One important question is the overall excess mortality of the observed population, irrespective of country. Figure S11 presents the fitting of the baseline mortality by age, adding together all the deaths and at-risk populations under analysis. Likewise, in the country-specific models, these baselines were fitted to annual deaths between 2015 and 2019 using a GLM model with overdispersion, and the prediction intervals were estimated through bootstrapping.

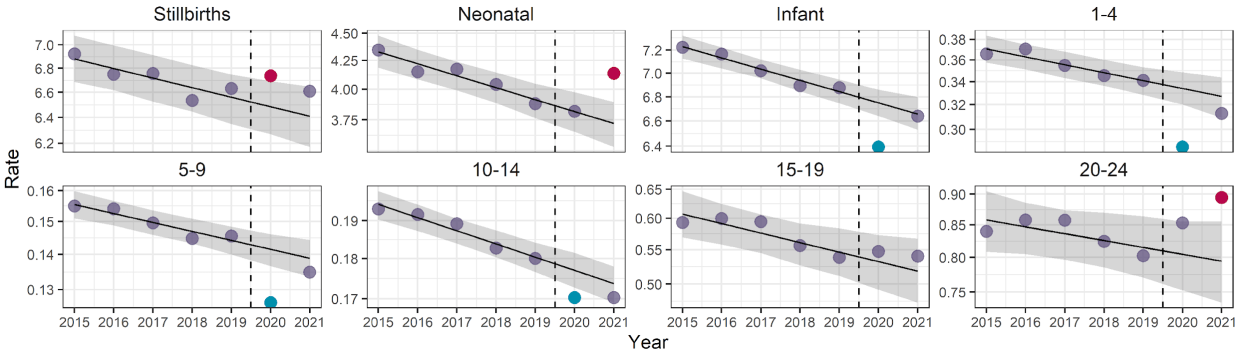

**Figure S11. Overall baseline mortality fitting by age group for all sexes combined, using a Generalized Linear Model with overdispersion.** Black dots indicate observed annual death counts between 2015 and 2019, and colored dots death counts in 2020 and 2021 indicate negative (deficit, in blue), positive (excess, in red), or no significant difference from expected mortality (in purple). The black line depicts the fitted baseline mortality, and the gray band, its bootstrapped 95% prediction interval.

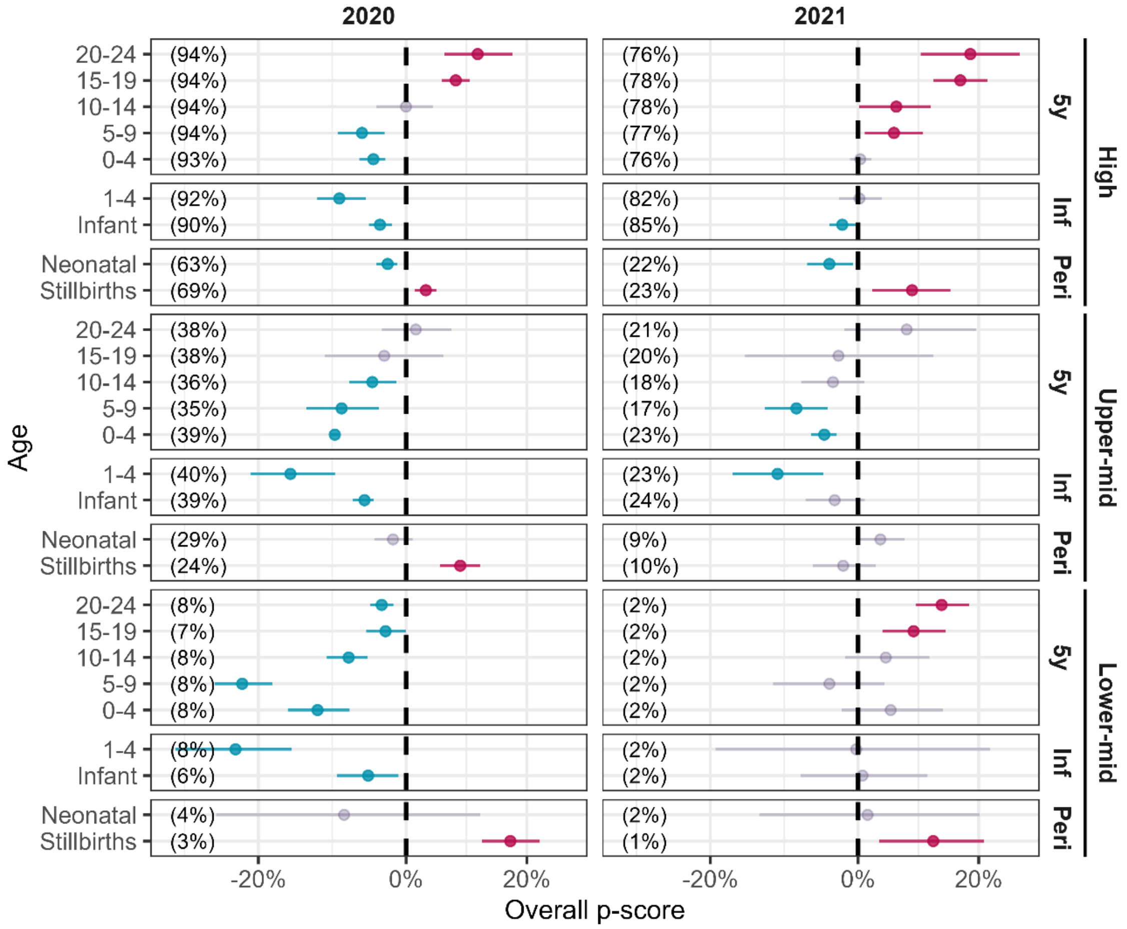

**Figure S12. Overall p-scores in 2020 and 2021 by age and income level.** Vertical dashed lines indicate no change in mortality relative to the expected value (i.e., p-score of 0%). Numbers in parentheses indicate the percentage of the total world population considered in the analysis within each age group. Points in blue and red indicate negative (deficit) and positive (excess) changes in mortality, respectively, that are statistically different from 0% ($\rho < 0.05$).

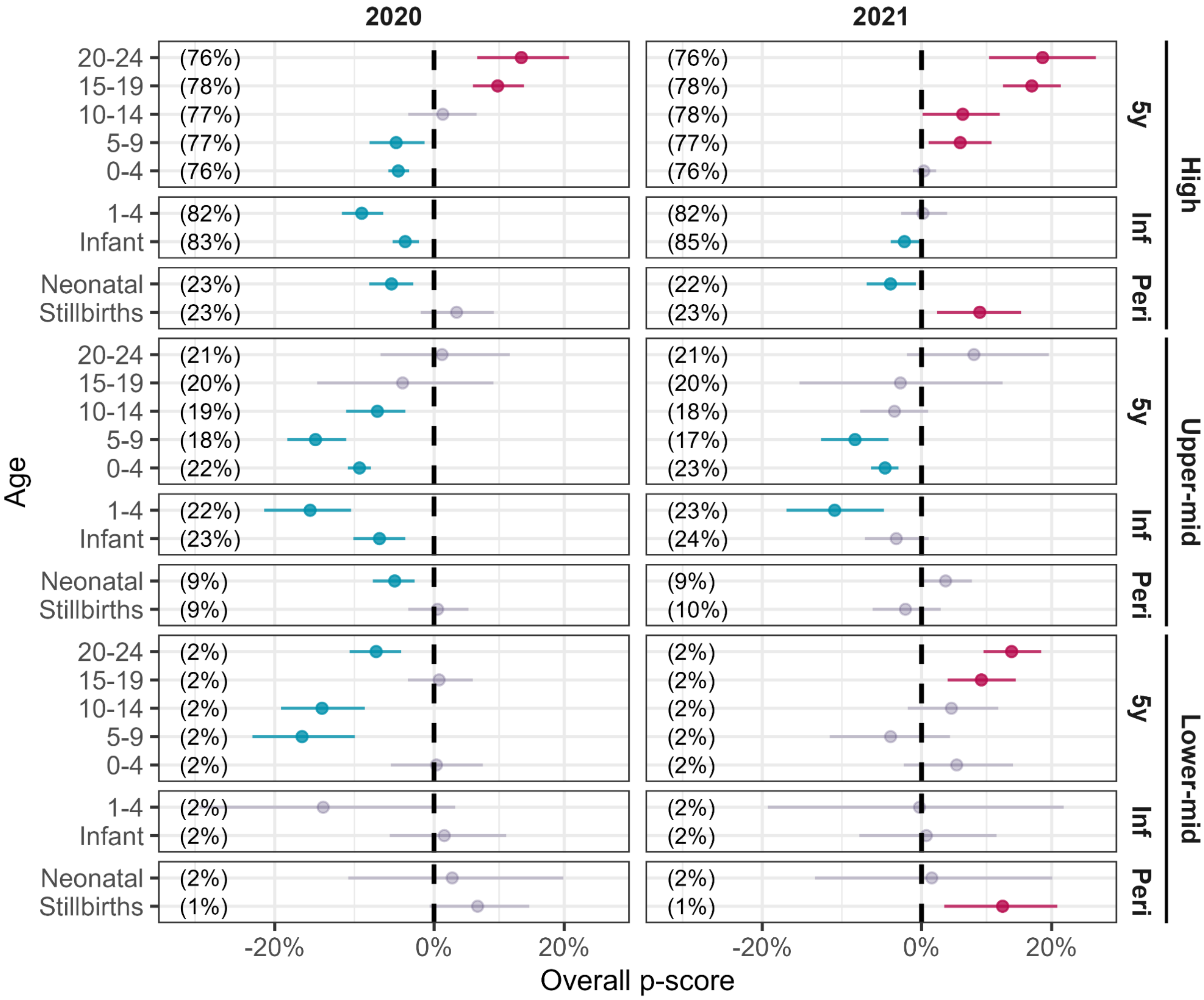

**Figure S13. Overall p-scores for the same countries in 2020 and 2021 by age and income level.** Vertical dashed lines indicate no change in mortality relative to the expected value (i.e., p-score of 0%). Numbers in parentheses indicate the percentage of the total world population considered in the analysis within each age group. Points in blue and red indicate negative (deficit) and positive (excess) changes in mortality, respectively, that are statistically different from 0% (ρ < 0.05).

### SENSITIVITY ANALYSES

#### Sensitivity test on population size

The p-score index, as any other relative measure, is highly sensitive to mortality changes in small populations. We included all available data, regardless of population size, to test how sensitive our estimates are to including small populations. Table S4 presents the number of countries and territories with available data and those where the fitting produced robust estimates.

**Table S4.** Countries and territories with available data and with robust baseline estimates on stillbirths and all-cause mortality by age and income level for years 2020 and 2021

| | | **Available data** | | **Robust Fitting** | |
|---|---|---|---|---|---|
| | | **2020** | **2021** | **2020** | **2021** |
| | **Total countries** | 111 | 71 | 104 | 68 |
| **Age at death** | Stillbirths (>28w) | 67 | 37 | 64 | 37 |
| | Neonatal (<28d) | 70 | 37 | 68 | 37 |
| | Infant (<1y) | 106 | 56 | 100 | 54 |
| | 1-4y | 104 | 55 | 91 | 50 |
| | 0-4y | 106 | 66 | 100 | 63 |
| | 5-9y | 106 | 66 | 85 | 54 |
| | 10-14y | 105 | 65 | 88 | 56 |
| | 15-19y | 103 | 64 | 92 | 57 |
| | 20-24y | 103 | 64 | 92 | 59 |
| **Income level** | Lower-middle | 13 | 5 | 13 | 5 |
| | Upper-middle | 35 | 20 | 34 | 20 |
| | High | 63 | 46 | 57 | 43 |

Using this threshold for population size, 27% (354 of 1,303) of age group-country combinations were excluded in this robustness check. Figure S13 depicts the weighted p-scores (circles) and the p-score values range (rhombus) for each age group in 2020 and 2021 when all countries are included (in black) and when countries with small populations are excluded (in red). As shown in **Fig. S14**, the truncation of combinations with small populations had an effect on the extreme values but not on the weighted average or the 25th and 75th p-score percentiles.

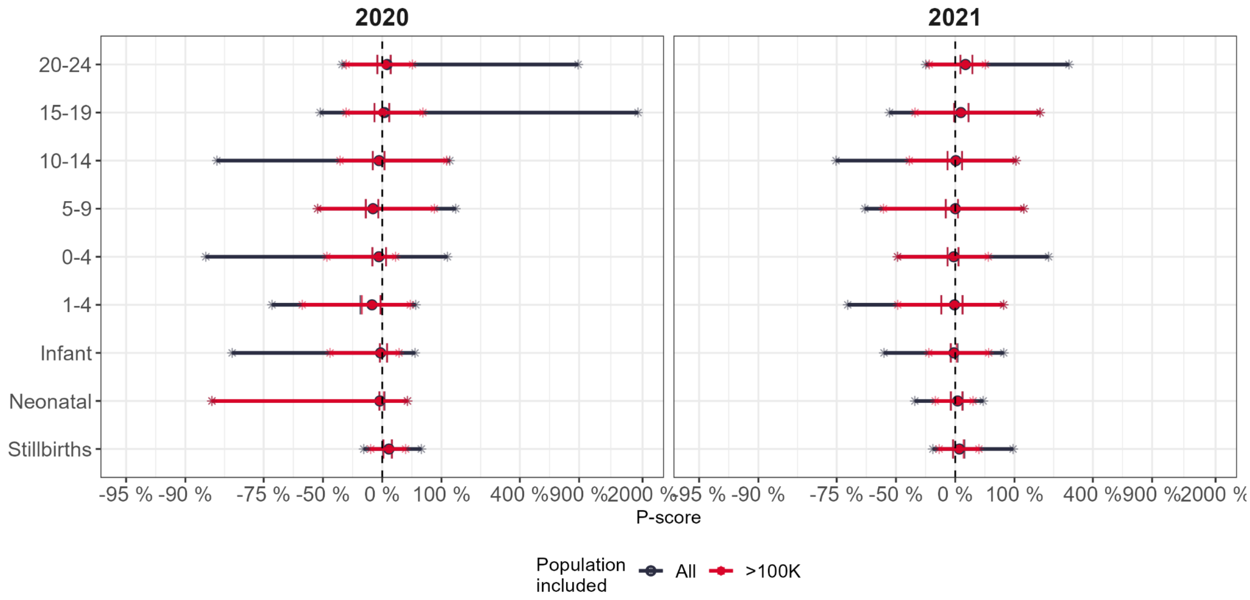

**Figure S14. Weighted p-score averages (circles) and p-score values range (asterisks) by age and year.** Circles indicate p-score averages in each age and year. Whiskers depict the 25th and 75th percentiles of p-score values in each age-year configuration. Summary measures of estimates that include all countries are depicted in black, and those excluding countries with small populations (<500K) are in red.

### Sensitivity test on fitting time resolution

Mortality data is available in weekly, monthly, and annual configurations. In this section, we test how sensitive excess estimates are to using different time-scale configurations in mortality data. Figure S15 presents the excess estimates obtained from weekly and annual mortality data. In both cases, we fit a Generalized Additive Model (GAM) with quasi-Poisson distribution to deaths between 2015 and 2019. Both models use a log-linear trend for the secular trend in mortality and the population at risk as an offset. The model for weekly data also includes a cyclic p-spline to account for within-year seasonality. Prediction intervals in both cases were estimated through bootstrapping with 1,000 iterations.

We find a negligible difference between excess estimates based on weekly or annual death counts in the countries where we have information in both time scales. On average, estimates based on annual deaths had a 0.07% lower p-score than estimates based on weekly deaths.

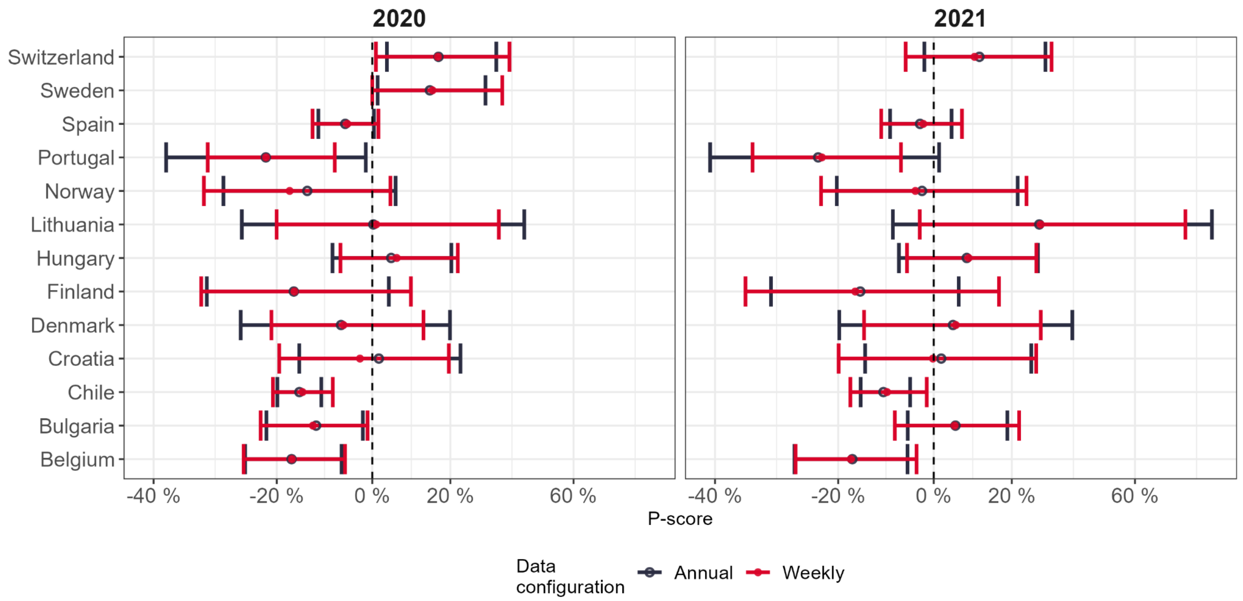

**Figure S15. Sensitivity analysis of p-score and its prediction interval according to different time scales.**

### Sensitivity test on previous years

This analysis allows us to evaluate whether the pattern in mortality changes observed in years 2020 and 2021 result from stochastic changes that would have been observed in any given year. For this purpose, we evaluate excess mortality estimates for 2017, 2018, and 2019. We estimate the baseline mortality based on the previous five years in each case (e.g., period 2012-2016 for estimating excess in 2017), using the same GLM model with quasi-Poisson distribution applied for estimating the 2020 and 2021 baselines. We then compute p-scores by country, age, and year and calculate the proportion of countries with negative and positive variations in mortality.

Figure S16 shows the distribution of p-scores for all years between 2017 and 2021, using the same set of **75** countries and territories for each year.

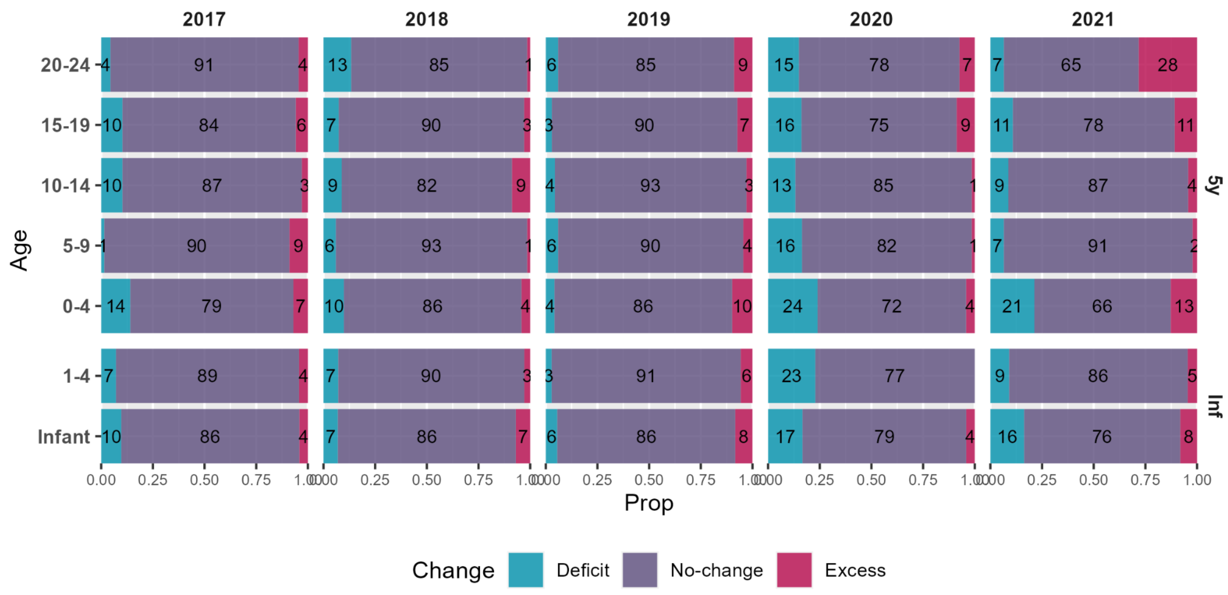

**Figure S16. Distribution of p-scores by population, age, and income level for all years between 2017 and 2021.** Vertical dashed lines indicate no change in mortality relative to the expected value (i.e., P-score = 0%). Bars in blue and red indicate the proportion of countries and territories with significant negative and positive changes in mortality, and purple bars indicate the proportion of countries and territories where changes were not statistically different from 0%.

Figure **S17** presents the age-specific overall p-score estimates for each year between 2017 and 2021 for the total population under observation and by income level. Figure **S18** shows these estimates but restricts the analysis to countries with data for all periods.

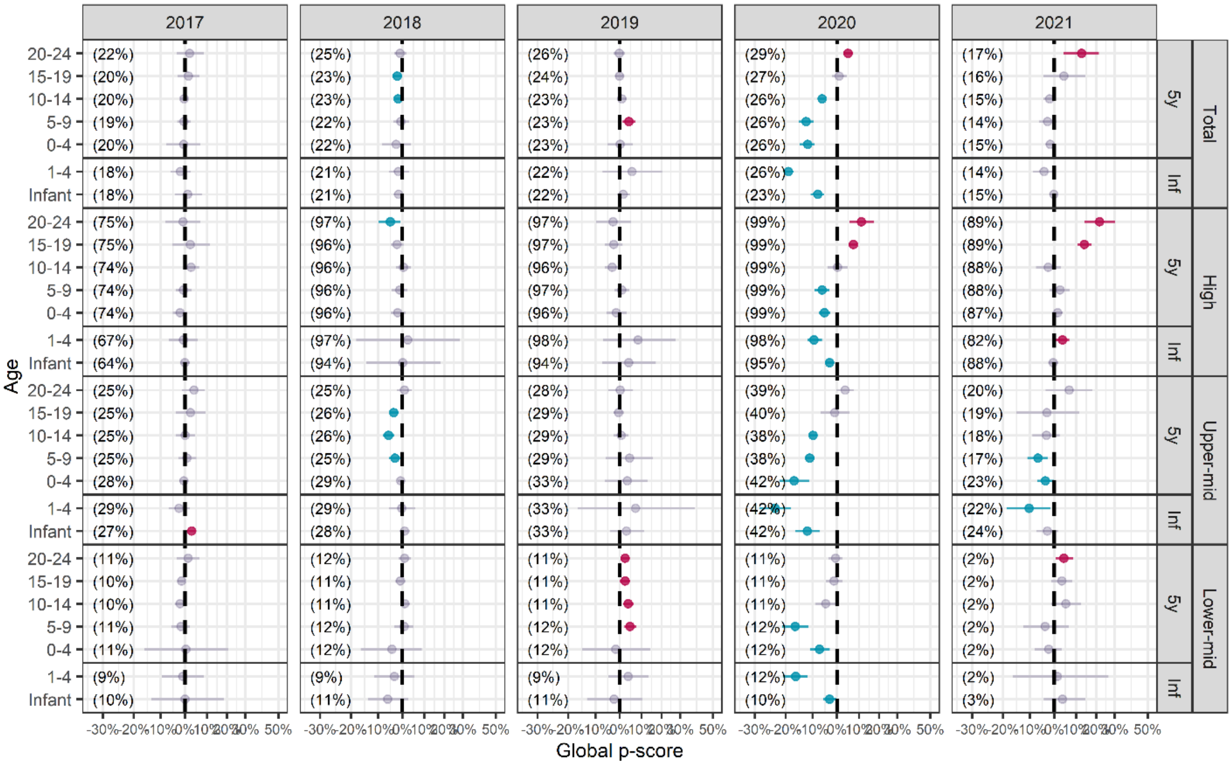

**Figure S18. Overall p-score estimates by age and income level for all years between 2017 and 2021.** Vertical dashed lines indicate no change in mortality relative to the expected value (i.e., P-score = 0%). Points in blue and red indicate negative (deficit) and positive (excess) changes in mortality, respectively, that are statistically different from 0% (ρ < 0.05), and gray points indicate changes within the prediction interval.

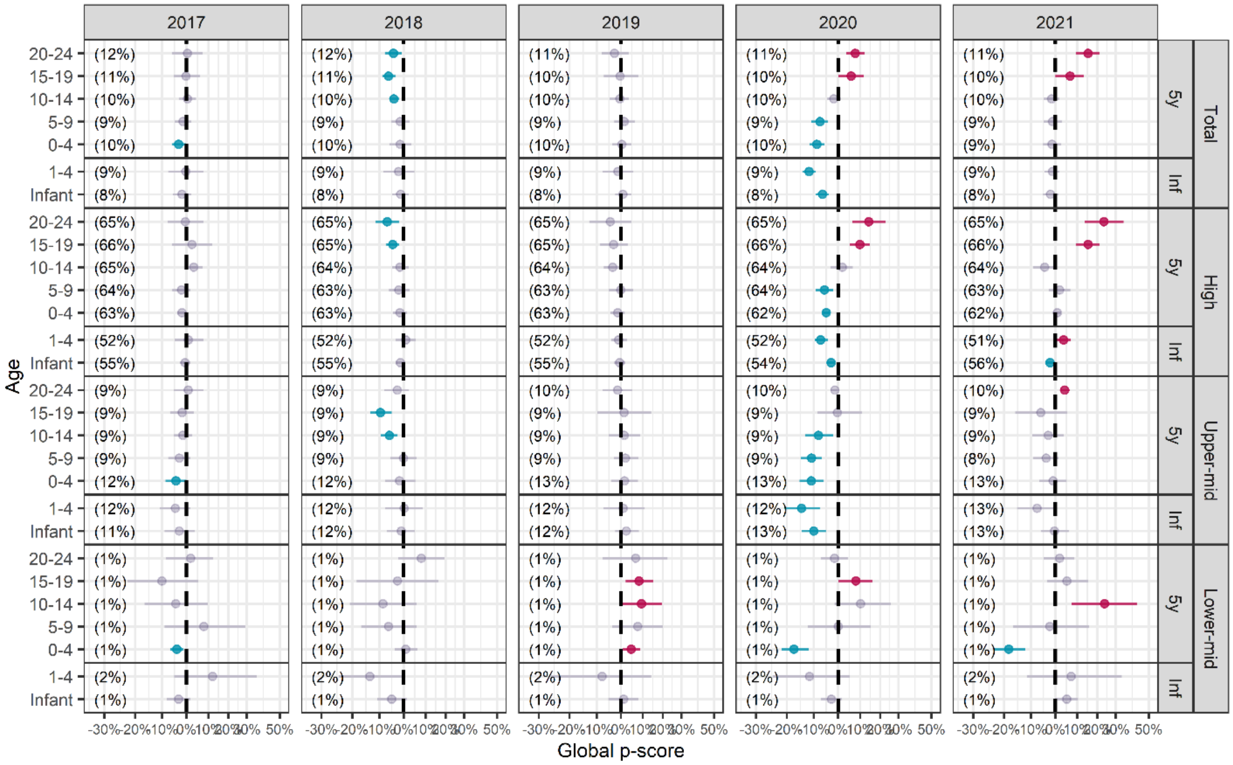

**Figure S19. Overall p-score estimates by age and income level for all years between 2017 and 2021.** Vertical dashed lines indicate no change in mortality relative to the expected value (i.e., P-score = 0%).Points in blue and red indicate negative (deficit) and positive (excess) changes in mortality, respectively, that are statistically different from 0% (ρ < 0.05) and gray points indicate changes within the prediction interval.

### TABLES WITH COUNTRY-SPECIFIC INFORMATION

#### Data availability by country

Table S3 presents detailed information for each country regarding the periods with available data on live births, stillbirths, and mortality among neonates, infants, and between ages 0 and 24.

**Table S3.** List of the countries and territories included in the analyses, indicating the ages observed and the years with available data on stillbirths and mortality of neonates, infants, and under 24y.

| Country | Live births | Stillbirths | Neonatal | Infant | Ages | Deaths |
|---|---|---|---|---|---|---|
| Afghanistan | 2017-2021 | 2017-2021 | - | - | 0-4 | 2017-2021 |
| Albania | 2010-2021 | 2017-2020 | 2013-2020 | 2013-2020 | 0-24 | 2015-2020 |
| American Samoa | 2010-2020 | - | - | 2010-2020 | 0-24 | 2010-2020 |
| Andorra | 2010-2021 | 2015-2020 | 2015-2020 | 2015-2020 | 0-24 | 2015-2020 |
| Antigua and Barbuda | 2010-2020 | - | - | 2012-2020 | 0-24 | 2012-2020 |
| Argentina | 2015-2020 | 2015-2020 | 2015-2020 | 2010-2020 | 0-24 | 2010-2020 |
| Aruba | 2010-2021 | - | - | 2012-2021 | 0-24 | 2012-2021 |
| Australia | 2010-2021 | 2015-2021 | 2015-2021 | 2010-2021 | 0-24 | 2010-2021 |
| Austria | 2010-2021 | 2011-2021 | 2011-2021 | 2010-2021 | 0-24 | 2010-2021 |
| Bangladesh | 2016-2021 | 2016-2021 | 2016-2021 | 2010-2020 | 0-4 | 2010-2020 |
| Belgium | 2010-2021 | 2011-2020 | 2011-2020 | 2010-2021 | 0-24 | 2010-2021 |
| Belize | 2010-2021 | - | - | 2015-2021 | 0-24 | 2015-2021 |
| Bermuda | 2010-2021 | - | - | 2010-2021 | 0-24 | 2010-2021 |
| Bosnia and Herzegovina | 2010-2021 | 2015-2020 | 2015-2020 | 2010-2020 | 0-24 | 2010-2020 |
| Brazil | 2010-2020 | 2015-2020 | 2015-2020 | 2015-2021 | 0-24 | 2015-2021 |
| Brunei Darussalam | 2010-2020 | - | - | 2013-2020 | 0-24 | 2013-2020 |
| Bulgaria | 2010-2021 | 2015-2021 | 2015-2021 | 2015-2021 | 0-24 | 2015-2021 |
| Burkina Faso | 2015-2021 | 2015-2021 | 2015-2021 | - | 0-4 | 2015-2021 |
| Burundi | 2018-2021 | 2018-2021 | 2018-2021 | - | - | - |
| Canada | 2010-2021 | 2015-2020 | 2015-2020 | 2015-2021 | 0-24 | 2015-2021 |
| Chile | 2010-2021 | - | - | 2015-2021 | 0-24 | 2015-2021 |
| China | - | - | - | 2010-2020 | 0-4 | 2010-2020 |
| Colombia | 2010-2021 | 2015-2021 | 2015-2021 | 2010-2021 | 0-24 | 2010-2021 |
| Costa Rica | 2010-2021 | - | 2010-2020 | 2015-2020 | 0-24 | 2015-2020 |
| Croatia | 2010-2021 | 2010-2020 | 2011-2020 | 2015-2021 | 0-24 | 2010-2021 |
| Cuba | 2010-2021 | - | 2010-2020 | 2010-2020 | 0-24 | 2010-2020 |
| Cyprus | 2010-2021 | 2015-2020 | 2015-2020 | 2010-2020 | 0-24 | 2015-2021 |
| Czechia | 2010-2021 | 2015-2021 | 2015-2021 | 2010-2021 | 0-24 | 2010-2021 |
| Denmark | 2010-2021 | 2011-2020 | 2011-2020 | 2010-2021 | 0-24 | 2010-2021 |
| Dominica | - | - | - | 2010-2020 | 0-24 | 2010-2020 |
| Dominican Republic | 2010-2021 | - | - | 2012-2021 | 0-24 | 2012-2021 |
| Ecuador | 2010-2021 | 2015-2021 | 2015-2021 | 2010-2021 | 0-24 | 2010-2021 |
| Egypt | 2010-2021 | - | - | 2010-2020 | 0-24 | 2010-2020 |
| Estonia | 2010-2021 | 2015-2020 | 2015-2020 | 2010-2021 | 0-24 | 2010-2021 |
| Eswatini | 2018-2021 | 2018-2021 | 2018-2021 | - | - | - |
| Ethiopia | 2018-2021 | 2018-2021 | 2018-2021 | - | 0-4 | 2018-2021 |
| Finland | 2010-2021 | 2015-2020 | 2015-2020 | 2010-2021 | 0-24 | 2010-2021 |
| France | 2010-2021 | 2015-2020 | 2011-2020 | 2010-2020 | 0-24 | 2013-2021 |
| French Guiana | 2010-2020 | - | - | 2013-2020 | 0-24 | 2013-2020 |
| French Polynesia | 2010-2021 | - | - | 2017-2021 | 0-24 | 2017-2021 |
| Georgia | 2010-2021 | 2015-2021 | 2015-2021 | 2010-2021 | 0-24 | 2010-2021 |
| Germany | 2010-2021 | 2011-2020 | 2011-2020 | 2010-2020 | 0-24 | 2010-2020 |
| Greece | 2010-2021 | 2015-2021 | 2015-2021 | 2014-2020 | 0-24 | 2015-2021 |
| Grenada | - | - | - | 2010-2021 | 0-24 | 2010-2021 |
| Guatemala | 2010-2020 | - | - | 2010-2020 | 0-24 | 2010-2020 |
| Honduras | 2015-2021 | 2015-2021 | - | 2010-2021 | 0-4 | 2010-2021 |
| Hong Kong - China SAR | - | - | - | 2010-2021 | 0-24 | 2010-2021 |

| Country | Live births | Stillbirths | Neonatal | Infant | Ages | Deaths |
|---|---|---|---|---|---|---|
| Hungary | 2010-2021 | 2015-2020 | 2015-2020 | 2010-2021 | 0-24 | 2010-2021 |
| Iceland | 2010-2021 | 2015-2020 | 2015-2020 | 2013-2021 | 0-24 | 2010-2021 |
| India | 2015-2021 | 2015-2021 | 2015-2021 | 2011-2020 | 0-4 | 2017-2021 |
| Iran | 2010-2020 | - | - | 2013-2020 | 0-24 | 2013-2020 |
| Ireland | 2010-2021 | 2011-2020 | 2011-2020 | 2010-2020 | 0-24 | 2010-2020 |
| Israel | 2010-2021 | 2015-2020 | 2015-2021 | 2010-2021 | 0-24 | 2010-2021 |
| Italy | 2010-2021 | 2011-2020 | 2011-2020 | 2011-2021 | 0-24 | 2011-2021 |
| Japan | 2010-2021 | 2015-2021 | 2015-2021 | 2010-2021 | 0-24 | 2010-2021 |
| Kazakhstan | 2010-2021 | - | 2010-2020 | 2010-2021 | 0-24 | 2010-2021 |
| Kenya | 2018-2021 | 2018-2021 | 2018-2021 | - | 0-4 | 2018-2022 |
| Kuwait | 2010-2021 | - | 2015-2020 | 2010-2020 | 0-24 | 2010-2020 |
| Kyrgyzstan | 2010-2021 | - | - | 2010-2020 | 0-24 | 2010-2020 |
| Latvia | 2010-2021 | 2015-2021 | 2015-2021 | 2010-2021 | 0-24 | 2010-2021 |
| Lebanon | - | - | - | 2017-2021 | 0-24 | 2017-2021 |
| Liberia | 2015-2021 | 2015-2021 | 2015-2021 | - | 0-4 | 2015-2021 |
| Libya | 2015-2020 | - | - | - | 5-24 | 2015-2020 |
| Liechtenstein | 2010-2021 | - | - | 2010-2020 | 0-24 | 2010-2021 |
| Lithuania | 2010-2021 | 2015-2021 | 2015-2021 | 2010-2021 | 0-24 | 2010-2021 |
| Luxembourg | 2010-2021 | 2011-2021 | 2011-2021 | 2010-2021 | 0-24 | 2010-2021 |
| Madagascar | 2018-2021 | 2018-2021 | 2018-2021 | - | - | - |
| Malawi | 2018-2021 | 2018-2021 | 2018-2020 | - | 0-4 | 2018-2020 |
| Malaysia | 2010-2021 | 2015-2021 | 2015-2021 | 2010-2021 | 0-24 | 2010-2021 |
| Maldives | 2010-2021 | 2015-2021 | 2015-2021 | 2010-2021 | 0-24 | 2010-2021 |
| Malta | 2010-2021 | 2015-2021 | 2015-2021 | 2010-2020 | 0-24 | 2011-2021 |
| Martinique | 2010-2020 | - | - | 2014-2020 | 0-24 | 2014-2020 |
| Mauritius | 2010-2021 | 2015-2021 | 2015-2021 | 2010-2021 | 0-24 | 2010-2021 |
| Mayotte | 2014-2020 | - | - | 2014-2020 | 0-24 | 2014-2020 |
| Mexico | 2010-2020 | 2015-2020 | 2015-2020 | 2010-2020 | 0-24 | 2010-2020 |
| Moldova | 2010-2021 | - | 2010-2020 | 2015-2021 | 0-24 | 2015-2021 |
| Mongolia | 2010-2021 | 2015-2021 | 2015-2021 | 2010-2021 | 0-24 | 2010-2021 |
| Montenegro | 2010-2021 | 2015-2021 | 2010-2020 | 2010-2020 | 0-24 | 2010-2021 |
| Montserrat | 2010-2021 | - | - | 2010-2021 | 0-24 | 2010-2021 |
| Mozambique | 2018-2021 | 2018-2021 | 2018-2021 | - | 0-4 | 2018-2021 |
| Nauru | 2015-2020 | 2015-2020 | 2015-2020 | 2015-2020 | 0-24 | 2015-2020 |
| Netherlands | 2010-2021 | 2011-2021 | 2011-2021 | 2010-2020 | 0-24 | 2010-2021 |
| New Zealand | 2010-2021 | - | - | 2010-2021 | 0-24 | 2010-2021 |
| Nicaragua | 2010-2020 | - | - | 2010-2020 | 0-24 | 2010-2020 |
| North Macedonia | 2010-2021 | 2010-2020 | 2010-2020 | 2010-2021 | 0-24 | 2010-2021 |
| Norway | 2010-2021 | 2015-2021 | 2015-2021 | 2010-2021 | 0-24 | 2010-2021 |
| Oman | 2010-2021 | 2015-2020 | 2015-2020 | 2010-2021 | 0-24 | 2010-2021 |
| Panama | 2010-2021 | 2015-2021 | 2015-2021 | 2010-2021 | 0-24 | 2010-2021 |
| Paraguay | - | - | - | 2010-2020 | 0-24 | 2010-2020 |
| Peru | 2010-2021 | - | - | 2010-2020 | 0-24 | 2010-2020 |
| Philippines | 2010-2021 | 2015-2020 | 2015-2020 | 2010-2020 | 0-24 | 2010-2020 |
| Poland | 2010-2021 | 2011-2021 | 2015-2021 | 2015-2021 | 0-24 | 2010-2021 |
| Portugal | 2010-2021 | 2015-2021 | 2015-2021 | 2010-2021 | 0-24 | 2010-2021 |
| Puerto Rico | 2010-2020 | - | - | 2013-2020 | 0-24 | 2013-2020 |
| Qatar | 2010-2021 | 2015-2020 | 2015-2020 | 2010-2020 | 0-24 | 2010-2020 |
| RÃ©union | 2010-2020 | - | - | 2013-2020 | 0-24 | 2013-2020 |
| Romania | 2010-2021 | 2015-2021 | 2015-2021 | 2015-2021 | 0-24 | 2015-2021 |
| Russia | 2010-2021 | - | 2015-2020 | 2015-2020 | 0-24 | 2010-2020 |
| Saint Lucia | - | - | - | 2010-2020 | 0-24 | 2010-2020 |
| Serbia | 2010-2021 | 2015-2021 | 2015-2021 | 2010-2021 | 0-24 | 2010-2021 |
| Seychelles | 2010-2021 | - | - | 2010-2021 | 0-24 | 2010-2021 |
| Singapore | 2010-2021 | 2015-2020 | 2015-2020 | 2010-2020 | 0-24 | 2010-2020 |
| Slovakia | 2010-2021 | 2015-2021 | 2015-2021 | 2010-2021 | 0-24 | 2010-2021 |
| Slovenia | 2010-2021 | 2015-2021 | 2015-2021 | 2010-2020 | 0-24 | 2010-2021 |
| South Africa | 2010-2020 | 2014-2021 | 2014-2021 | 2010-2021 | 1-24 | 2010-2021 |
| South Korea | 2010-2021 | 2015-2021 | 2015-2021 | 2015-2021 | 0-24 | 2010-2020 |
| Spain | 2010-2021 | 2011-2021 | 2011-2021 | 2010-2021 | 0-24 | 2010-2021 |
| Sri Lanka | 2010-2021 | 2017-2021 | 2017-2021 | 2010-2021 | 0-14 | 2010-2021 |
| State of Palestine | 2010-2020 | - | - | 2012-2020 | 0-24 | 2012-2020 |
| Suriname | 2010-2020 | - | - | 2012-2020 | 0-24 | 2012-2020 |

| Country | Live births | Stillbirths | Neonatal | Infant | Ages | Deaths |
|---|---|---|---|---|---|---|
| Sweden | 2010-2021 | 2015-2020 | 2015-2020 | 2010-2021 | 0-24 | 2010-2021 |
| Switzerland | 2010-2021 | 2015-2021 | 2015-2021 | 2010-2021 | 0-24 | 2010-2021 |
| Taiwan - Province of China | 2010-2021 | - | - | - | 0-24 | 2010-2021 |
| Thailand | 2010-2021 | - | - | 2010-2021 | 0-24 | 2010-2021 |
| Turkey | 2010-2021 | 2015-2021 | 2015-2021 | 2010-2021 | 0-4 | 2010-2021 |
| Turks and Caicos Islands | 2010-2020 | - | - | 2014-2020 | 0-24 | 2014-2020 |
| Uganda | 2018-2021 | 2018-2021 | 2018-2021 | - | 0-4 | 2018-2022 |
| Ukraine | 2010-2021 | 2015-2021 | 2015-2021 | 2010-2021 | 0-24 | 2010-2021 |
| United Arab Emirates | 2010-2020 | - | - | 2016-2020 | 0-24 | 2016-2020 |
| United Kingdom | 2010-2020 | 2015-2020 | 2015-2020 | 2010-2020 | 0-24 | 2010-2020 |
| Uruguay | 2010-2021 | 2015-2021 | 2015-2021 | 2010-2021 | 0-24 | 2010-2021 |
| USA | 2010-2021 | 2015-2020 | 2015-2020 | 2010-2021 | 0-24 | 2010-2021 |
| Uzbekistan | 2010-2021 | - | 2015-2021 | 2010-2021 | 0-24 | 2010-2021 |
| Zambia | 2018-2021 | 2018-2021 | 2018-2021 | - | - | - |
| Zimbabwe | 2018-2021 | 2018-2021 | 2018-2021 | - | - | - |

#### Data sources by country

Table S4 presents the sources in each country from which data on live births, stillbirths, neonatal, infant, and under-25 mortality was obtained.

**Table S4.** List of the populations included in the analyses, indicating the sources of information on stillbirths and mortality of neonates, infants, and under 24y.

| Country / Territory | Measure | Source |
|---|---|---|
| Afghanistan | live births | Health Management Information System (HMIS) |
| Albania | live births | UNPD - Demographic Yearbook |
| American Samoa | live births | UNPD - Demographic Yearbook |
| Andorra | live births | UNPD - Demographic Yearbook |
| Antigua and Barbuda | live births | UNPD - Demographic Yearbook |
| Argentina | live births | UNICEF data call. Argentina Ministerio de Salud - CRVS. Data submitted to UNICEF via e-mail |
| Aruba | live births | UNPD - Demographic Yearbook |
| Australia | live births | UNICEF data call. Australian Bureau of Statistics. Vital statistics and civil registration (CRVS). https://www. abs.gov.au. |
| Austria | live births | Short-term fertility fluctuations database (STFF) |
| Bangladesh | live births | Health Management Information System (HMIS) |
| Belgium | live births | Human Mortality Database (HMD) |
| Belize | live births | UNPD - Demographic Yearbook |
| Bermuda | live births | UNPD - Demographic Yearbook |
| Bosnia and Herzegovina | live births | UNPD - Demographic Yearbook |
| Brazil | live births | Ministerio da Saude - Datasus https://datasus.saude.gov.br/ |
| Brunei Darussalam | live births | UNPD - Demographic Yearbook |
| Bulgaria | live births | UNICEF data call. National Statistical Institute of Bulgaria, Information System Demography. https://www.czso.cz/csu/czso/home. |
| Burkina Faso | live births | Health Management Information System (HMIS) |
| Burundi | live births | Health Management Information System (HMIS) |
| Canada | live births | Short-term fertility fluctuations database (STFF) |
| Chile | live births | Short-term fertility fluctuations database (STFF) |
| China | live births | UNPD - World Population Pospects 2022 (WPP) |
| Colombia | live births | UNICEF data call. National Administrative Department of Statistics (DANE) - CRVS. Data submitted to UNICEF via e-mail |
| Costa Rica | live births | UNPD - Demographic Yearbook |
| Croatia | live births | Eurostat Database |
| Cuba | live births | UNPD - Demographic Yearbook |
| Cyprus | live births | Eurostat Database |
| Czechia | live births | UNICEF data call. Czech Statistical Office. CRVS-statistical reports on birth/death. https://www.czso.cz/csu/czso/home. |
| Denmark | live births | Human Mortality Database (HMD) |

| Country / Territory | Measure | Source |
|---|---|---|
| Dominica | live births | UNPD - World Population Pospects 2022 (WPP) |
| Dominican Republic | live births | UNPD - Demographic Yearbook |
| Ecuador | live births | UNICEF data call. Instituto Nacional de EstadÃƒÂstica y Censos (INEC) - CRVS. Data submitted to UNICEF via e-mail |
| Egypt | live births | UNPD - Demographic Yearbook |
| Estonia | live births | Short-term fertility fluctuations database (STFF) |
| Eswatini | live births | Health Management Information System (HMIS) |
| Ethiopia | live births | Health Management Information System (HMIS) |
| Finland | live births | UNICEF data call. Statistics Finland. Digital and Population Data Services Agency. |
| France | live births | Eurostat Database |
| French Guiana | live births | UNPD - Demographic Yearbook |
| French Polynesia | live births | UNPD - Demographic Yearbook |
| Georgia | live births | UNICEF data call. National Statistics Office of Georgia (Geostat). Administrative data. |
| Germany | live births | Eurostat Database |
| Greece | live births | Eurostat Database |
| Grenada | live births | UNPD - World Population Pospects 2022 (WPP) |
| Guatemala | live births | UNPD - Demographic Yearbook |
| Honduras | live births | UNICEF data call. Secretaria de Salud of Honduras. Area Estadistica de la Salud. Data submitted to UNICEF via e-mail |
| Hong Kong - China SAR | live births | UNPD - World Population Pospects 2022 (WPP) |
| Hungary | live births | Eurostat Database |
| Iceland | live births | Short-term fertility fluctuations database (STFF) |
| India | live births | Ministry of Health and Family Welfare of India. Health Management Information System (HMIS). https://hmis.mohfw.gov.in |
| Iran | live births | UNPD - Demographic Yearbook |
| Ireland | live births | Eurostat Database |
| Israel | live births | Short-term fertility fluctuations database (STFF) |
| Italy | live births | Eurostat Database |
| Japan | live births | Human Mortality Database (HMD) |
| Kazakhstan | live births | UNPD - Demographic Yearbook |
| Kenya | live births | Health Management Information System (HMIS) |
| Kuwait | live births | UNPD - Demographic Yearbook |
| Kyrgyzstan | live births | UNPD - Demographic Yearbook |
| Latvia | live births | Short-term fertility fluctuations database (STFF) |
| Lebanon | live births | UNPD - World Population Pospects 2022 (WPP) |
| Liberia | live births | Health Management Information System (HMIS) |
| Libya | live births | UNICEF data call. Libyan Civil Registration Authority. Electronic system based on actual deaths and births registered by date. |
| Liechtenstein | live births | Eurostat Database |
| Lithuania | live births | UNICEF data call. Statistics Lithuania. Population register. |
| Luxembourg | live births | Human Mortality Database (HMD) |
| Madagascar | live births | Health Management Information System (HMIS) |
| Malawi | live births | Health Management Information System (HMIS) |
| Malaysia | live births | UNICEF data call. Ministry of Health of Malaysia. Department of National Registration. |
| Maldives | live births | UNICEF data call. Maldives Ministry of Health. Vital registration. |
| Malta | live births | UNICEF data call. Ministry for Health and National Statistics Office of Malta. Malta National Birth and Mortality Registry. Data submitted to UNICEF via email. |
| Martinique | live births | UNPD - Demographic Yearbook |
| Mauritius | live births | UNICEF data call. Statistics Mauritius, Vital statistics and civil registration. |
| Mayotte | live births | UNPD - Demographic Yearbook |
| Mexico | live births | Instituto Nacional de Estadistica y Geografia (INEGI) - CRVS |
| Moldova | live births | UNPD - Demographic Yearbook |
| Mongolia | live births | UNPD - Demographic Yearbook |
| Montenegro | live births | UNICEF data call. Montenegro Statistical Office and Institute of Public Health of Montenegro. Vital registration data. Data submitted to UNICEF via email. |
| Montserrat | live births | UNPD - Demographic Yearbook |
| Mozambique | live births | Countrywide Mortality Surveillance for Action (COMSA) |
| Nauru | live births | UNICEF data call. Government of the Republic of Nauru. Birth Notification and Certificate and Cause of Death Notifification. |
| Netherlands | live births | Short-term fertility fluctuations database (STFF) |
| New Zealand | live births | UNPD - Demographic Yearbook |
| Nicaragua | live births | UNPD - Demographic Yearbook |
| North Macedonia | live births | UNPD - Demographic Yearbook |
| Norway | live births | Human Mortality Database (HMD) |

| Country / Territory | Measure | Source |
|---|---|---|
| Oman | live births | UNPD - Demographic Yearbook |
| Panama | live births | UNICEF data call. Panama Ministry of Health. Vital statistics and civil registration. |
| Paraguay | live births | UNPD - World Population Pospects 2022 (WPP) |
| Peru | live births | UNPD - Demographic Yearbook |
| Philippines | live births | UNPD - Demographic Yearbook |
| Poland | live births | UNICEF data call. Statisitics Poland. Civil Registration and Vital Statistics. https://stat.gov.pl/. |
| Portugal | live births | UNICEF data call. Statistics Portugal. Civil Registration and Vital Statistics. |
| Puerto Rico | live births | UNPD - Demographic Yearbook |
| Qatar | live births | UNPD - Demographic Yearbook |
| Reunion | live births | UNPD - Demographic Yearbook |
| Romania | live births | Short-term fertility fluctuations database (STFF) |
| Russia | live births | UNPD - Demographic Yearbook |
| Saint Lucia | live births | UNPD - World Population Pospects 2022 (WPP) |
| Serbia | live births | UNPD - Demographic Yearbook |
| Seychelles | live births | UNPD - Demographic Yearbook |
| Singapore | live births | UNPD - Demographic Yearbook |
| Slovakia | live births | Short-term fertility fluctuations database (STFF) |
| Slovenia | live births | Short-term fertility fluctuations database (STFF) |
| South Africa | live births | UNPD - Dorrington, R. |
| South Korea | live births | UNICEF data call. Statisitcs Korea. Civil registration system. https://kosis.kr, https://mdis.kostat.go.kr. |
| Spain | live births | Short-term fertility fluctuations database (STFF) |
| Sri Lanka | live births | UNPD - Demographic Yearbook |
| State of Palestine | live births | UNPD - Demographic Yearbook |
| Suriname | live births | UNPD - Demographic Yearbook |
| Sweden | live births | Human Mortality Database (HMD) |
| Switzerland | live births | UNICEF data call. Federal statistical office, Switzerland. Vita Statistics, BEVNAT. https://www.bfs.admin.ch/bfs/en/home/statistics/population/births-deaths/. |
| Taiwan - Province of China | live births | Short-term fertility fluctuations database (STFF) |
| Thailand | live births | UNPD - Demographic Yearbook |
| Turkey | live births | UNICEF data call. Turkish Statistical Institute (TURKSTAT) and Turkish Minsitry of Health. Central Civil Registration System (MERNÃ„Â°S) and Death Notification System (OBS). |
| Turks and Caicos Islands | live births | UNPD - Demographic Yearbook |
| Uganda | live births | Health Management Information System (HMIS) |
| Ukraine | live births | Short-term fertility fluctuations database (STFF) |
| United Arab Emirates | live births | UNPD - Demographic Yearbook |
| United Kingdom | live births | Human Mortality Database (HMD) |
| Uruguay | live births | UNICEF data call. Uruguay Ministry of Health. Vital Statistics. https://uins.msp.gub.uy/. |
| USA | live births | Human Mortality Database (HMD) |
| Uzbekistan | live births | UNPD - Demographic Yearbook |
| Zambia | live births | Health Management Information System (HMIS) |
| Zimbabwe | live births | Health Management Information System (HMIS) |
| Albania | population | UNPD - World Population Pospects 2022 (WPP) |
| American Samoa | population | UNPD - World Population Pospects 2022 (WPP) |
| Andorra | population | UNPD - World Population Pospects 2022 (WPP) |
| Antigua and Barbuda | population | UNPD - World Population Pospects 2022 (WPP) |
| Argentina | population | UNPD - World Population Pospects 2022 (WPP) |
| Aruba | population | UNPD - World Population Pospects 2022 (WPP) |
| Australia | population | Human Mortality Database (HMD) |
| Austria | population | UNPD - World Population Pospects 2022 (WPP) |
| Bangladesh | population | UNPD - World Population Pospects 2022 (WPP) |
| Belgium | population | Human Mortality Database (HMD) |
| Belize | population | UNPD - World Population Pospects 2022 (WPP) |
| Bermuda | population | UNPD - World Population Pospects 2022 (WPP) |
| Bosnia and Herzegovina | population | UNPD - World Population Pospects 2022 (WPP) |
| Brazil | population | UNPD - World Population Pospects 2022 (WPP) |
| Brunei Darussalam | population | UNPD - World Population Pospects 2022 (WPP) |
| Bulgaria | population | Human Mortality Database (HMD) |
| Canada | population | Human Mortality Database (HMD) |

| Country / Territory | Measure | Source |
|---|---|---|
| Chile | population | Human Mortality Database (HMD) |
| China | population | UNPD - World Population Pospects 2022 (WPP) |
| Colombia | population | UNPD - World Population Pospects 2022 (WPP) |
| Costa Rica | population | UNPD - World Population Pospects 2022 (WPP) |
| Croatia | population | Human Mortality Database (HMD) |
| Cuba | population | UNPD - World Population Pospects 2022 (WPP) |
| Cyprus | population | UNPD - World Population Pospects 2022 (WPP) |
| Czechia | population | Human Mortality Database (HMD) |
| Denmark | population | Human Mortality Database (HMD) |
| Dominica | population | UNPD - World Population Pospects 2022 (WPP) |
| Dominican Republic | population | UNPD - World Population Pospects 2022 (WPP) |
| Ecuador | population | UNPD - World Population Pospects 2022 (WPP) |
| Egypt | population | UNPD - World Population Pospects 2022 (WPP) |
| Estonia | population | UNPD - World Population Pospects 2022 (WPP) |
| Finland | population | Human Mortality Database (HMD) |
| France | population | Human Mortality Database (HMD) |
| French Guiana | population | UNPD - World Population Pospects 2022 (WPP) |
| French Polynesia | population | UNPD - World Population Pospects 2022 (WPP) |
| Georgia | population | UNPD - World Population Pospects 2022 (WPP) |
| Germany | population | Human Mortality Database (HMD) |
| Greece | population | UNPD - World Population Pospects 2022 (WPP) |
| Grenada | population | UNPD - World Population Pospects 2022 (WPP) |
| Guatemala | population | UNPD - World Population Pospects 2022 (WPP) |
| Honduras | population | UNPD - World Population Pospects 2022 (WPP) |
| Hong Kong - China SAR | population | UNPD - World Population Pospects 2022 (WPP) |
| Hungary | population | Human Mortality Database (HMD) |
| Iceland | population | Human Mortality Database (HMD) |
| India | population | UNPD - World Population Pospects 2022 (WPP) |
| Iran | population | UNPD - World Population Pospects 2022 (WPP) |
| Ireland | population | Human Mortality Database (HMD) |
| Israel | population | UNPD - World Population Pospects 2022 (WPP) |
| Italy | population | UNPD - World Population Pospects 2022 (WPP) |
| Japan | population | Human Mortality Database (HMD) |
| Kazakhstan | population | UNPD - World Population Pospects 2022 (WPP) |
| Kuwait | population | UNPD - World Population Pospects 2022 (WPP) |
| Kyrgyzstan | population | UNPD - World Population Pospects 2022 (WPP) |
| Latvia | population | UNPD - World Population Pospects 2022 (WPP) |
| Lebanon | population | UNPD - World Population Pospects 2022 (WPP) |
| Libya | population | UNPD - World Population Pospects 2022 (WPP) |
| Liechtenstein | population | UNPD - World Population Pospects 2022 (WPP) |
| Lithuania | population | Human Mortality Database (HMD) |
| Luxembourg | population | Human Mortality Database (HMD) |
| Malaysia | population | UNPD - World Population Pospects 2022 (WPP) |
| Maldives | population | UNPD - World Population Pospects 2022 (WPP) |
| Malta | population | UNPD - World Population Pospects 2022 (WPP) |
| Martinique | population | UNPD - World Population Pospects 2022 (WPP) |
| Mauritius | population | UNPD - World Population Pospects 2022 (WPP) |
| Mayotte | population | UNPD - World Population Pospects 2022 (WPP) |
| Mexico | population | UNPD - World Population Pospects 2022 (WPP) |
| Moldova | population | UNPD - World Population Pospects 2022 (WPP) |
| Mongolia | population | UNPD - World Population Pospects 2022 (WPP) |
| Montenegro | population | UNPD - World Population Pospects 2022 (WPP) |
| Montserrat | population | UNPD - World Population Pospects 2022 (WPP) |
| Nauru | population | UNPD - World Population Pospects 2022 (WPP) |
| Netherlands | population | UNPD - World Population Pospects 2022 (WPP) |
| New Zealand | population | Human Mortality Database (HMD) |
| Nicaragua | population | UNPD - World Population Pospects 2022 (WPP) |
| North Macedonia | population | UNPD - World Population Pospects 2022 (WPP) |
| Norway | population | Human Mortality Database (HMD) |
| Oman | population | UNPD - World Population Pospects 2022 (WPP) |
| Panama | population | UNPD - World Population Pospects 2022 (WPP) |
| Paraguay | population | UNPD - World Population Pospects 2022 (WPP) |
| Peru | population | UNPD - World Population Pospects 2022 (WPP) |

| Country / Territory | Measure | Source |
|---|---|---|
| Philippines | population | UNPD - World Population Pospects 2022 (WPP) |
| Poland | population | UNPD - World Population Pospects 2022 (WPP) |
| Portugal | population | Human Mortality Database (HMD) |
| Puerto Rico | population | UNPD - World Population Pospects 2022 (WPP) |
| Qatar | population | UNPD - World Population Pospects 2022 (WPP) |
| Reunion | population | UNPD - World Population Pospects 2022 (WPP) |
| Romania | population | UNPD - World Population Pospects 2022 (WPP) |
| Russia | population | UNPD - World Population Pospects 2022 (WPP) |
| Saint Lucia | population | UNPD - World Population Pospects 2022 (WPP) |
| Serbia | population | UNPD - World Population Pospects 2022 (WPP) |
| Seychelles | population | UNPD - World Population Pospects 2022 (WPP) |
| Singapore | population | UNPD - World Population Pospects 2022 (WPP) |
| Slovakia | population | UNPD - World Population Pospects 2022 (WPP) |
| Slovenia | population | UNPD - World Population Pospects 2022 (WPP) |
| South Africa | population | UNPD - World Population Pospects 2022 (WPP) |
| South Korea | population | Human Mortality Database (HMD) |
| Spain | population | Human Mortality Database (HMD) |
| Sri Lanka | population | UNPD - World Population Pospects 2022 (WPP) |
| State of Palestine | population | UNPD - World Population Pospects 2022 (WPP) |
| Suriname | population | UNPD - World Population Pospects 2022 (WPP) |
| Sweden | population | Human Mortality Database (HMD) |
| Switzerland | population | Human Mortality Database (HMD) |
| Taiwan - Province of China | population | UNPD - World Population Pospects 2022 (WPP) |
| Thailand | population | UNPD - World Population Pospects 2022 (WPP) |
| Turkey | population | UNPD - World Population Pospects 2022 (WPP) |
| Turks and Caicos Islands | population | UNPD - World Population Pospects 2022 (WPP) |
| Ukraine | population | UNPD - World Population Pospects 2022 (WPP) |
| United Arab Emirates | population | UNPD - World Population Pospects 2022 (WPP) |
| United Kingdom | population | Human Mortality Database (HMD) |
| Uruguay | population | UNPD - World Population Pospects 2022 (WPP) |
| USA | population | Human Mortality Database (HMD) |
| Uzbekistan | population | UNPD - World Population Pospects 2022 (WPP) |
| Afghanistan | stillbirths | Health Management Information System (HMIS) |
| Albania | stillbirths | Eurostat Database |
| Andorra | stillbirths | UNICEF data call. Andorra Ministry of Health. Mortality and birth register of https://www.salut.ad/temes-de-salut/natalitat-i-mortalitat. |
| Argentina | stillbirths | UNICEF data call. Argentina Ministerio de Salud - CRVS. Data submitted to UNICEF via e-mail |
| Australia | stillbirths | UNICEF data call. Australian Bureau of Statistics. Vital statistics and civil registration (CRVS). https://www. abs.gov.au. |
| Austria | stillbirths | Eurostat Database |
| Bangladesh | stillbirths | Health Management Information System (HMIS) |
| Belgium | stillbirths | Eurostat Database |
| Bosnia and Herzegovina | stillbirths | UNICEF data call. Agency for Statistics of Bosnia and Herzegovina. Civil registration and vital statistics. |
| Brazil | stillbirths | UNICEF data call. Ministerio da Saude - Datasus https://datasus.saude.gov.br/ |
| Bulgaria | stillbirths | UNICEF data call. National Statistical Institute of Bulgaria, Information System Demography. https://www.czso.cz/csu/czso/home. |
| Burkina Faso | stillbirths | Health Management Information System (HMIS) |
| Burundi | stillbirths | Health Management Information System (HMIS) |
| Canada | stillbirths | UNICEF data call. Statistics Canada. Vital statistics and civil registration (CRVS). https://www23.statcan.gc.ca/ |
| Colombia | stillbirths | National Administrative Department of Statistics (DANE) - CRVS |
| Croatia | stillbirths | Eurostat Database |
| Cyprus | stillbirths | UNICEF data call. Republic of Cyprus. Ministry of Health. Medical Birth and Death Registry. |
| Czechia | stillbirths | UNICEF data call. Czech Statistical Office. CRVS-statistical reports on birth/death. https://www.czso.cz/csu/czso/home. |
| Denmark | stillbirths | Eurostat Database |
| Ecuador | stillbirths | UNICEF data call. Instituto Nacional de EstadÃƒÂstica y Censos (INEC) - CRVS. Data submitted to UNICEF via e-mail |

| Country / Territory | Measure | Source |
|---|---|---|
| Estonia | stillbirths | UNICEF data call. National Institute for Health Development, Estonia. Estonian Medical Birth Registry and Estonian Abortion Registry and Estonian Causes of Death Registry |
| Eswatini | stillbirths | Health Management Information System (HMIS) |
| Ethiopia | stillbirths | Health Management Information System (HMIS) |
| Finland | stillbirths | UNICEF data call. Statistics Finland. Digital and Population Data Services Agency. |
| France | stillbirths | UNICEF data call. SystÃƒÂ¨me National des DonnÃƒÂ©es de SantÃƒÂ© (SNDS) - CRVS |
| Georgia | stillbirths | UNICEF data call. National Statistics Office of Georgia (Geostat). |
| Germany | stillbirths | Eurostat Database |
| Greece | stillbirths | UNICEF data call. Hellenic Statistical Authority - CRVS. Data submitted to UNICEF via email. |
| Honduras | stillbirths | UNICEF data call. Secretaria de Salud of Honduras. Area Estadistica de la Salud. Data submitted to UNICEF via e-mail |
| Hungary | stillbirths | UNICEF data call. Hungarian Central Statistical Office. Department of Population Statistics. https://www.ksh.hu/. |
| Iceland | stillbirths | UNICEF data call. Icelandic Medical Birth Registry |
| India | stillbirths | Ministry of Health and Family Welfare of India. Health Management Information System (HMIS). https://hmis.mohfw.gov.in |
| Ireland | stillbirths | Eurostat Database |
| Israel | stillbirths | UNICEF data call. Israel Central Bureau of Statistics. Forms from hospitals for stillbirths. www.cbs.gov.il. |
| Italy | stillbirths | Eurostat Database |
| Japan | stillbirths | UNICEF data call. Ministry of Health of Japan, Labour and Welfare. Vital Statistics of Japan. https://www.e-stat.go.jp/. |
| Kenya | stillbirths | Health Management Information System (HMIS) |
| Latvia | stillbirths | UNICEF data call. Latvia Ministry of Health, The Centre for Disease Prevention and Control of Latvia. Medical Birth Register and Register of Causes of Death. |
| Liberia | stillbirths | Health Management Information System (HMIS) |
| Lithuania | stillbirths | UNICEF data call. Statistics Lithuania. Population register. |
| Luxembourg | stillbirths | Eurostat Database |
| Madagascar | stillbirths | Health Management Information System (HMIS) |
| Malawi | stillbirths | Health Management Information System (HMIS) |
| Malaysia | stillbirths | UNICEF data call. Ministry of Health of Malaysia. Department of National Registration. |
| Maldives | stillbirths | UNICEF data call. Maldives Ministry of Health. Vital registration. |
| Malta | stillbirths | UNICEF data call. Ministry for Health and National Statistics Office of Malta. Malta National Birth and Mortality Registry. Data submitted to UNICEF via email. |
| Mauritius | stillbirths | UNICEF data call. Statistics Mauritius, Vital statistics and civil registration. |
| Mexico | stillbirths | Instituto Nacional de Estadistica y Geografia (INEGI) - CRVS |
| Mongolia | stillbirths | UNICEF data call. Mongolia Ministry of Health. National Center for Health Development. Administrative records. |
| Montenegro | stillbirths | UNICEF data call. Montenegro Statistical Office and Institute of Public Health of Montenegro. Vital registration data. Data submitted to UNICEF via email. |
| Mozambique | stillbirths | Countrywide Mortality Surveillance for Action (COMSA) |
| Nauru | stillbirths | UNICEF data call. Government of the Republic of Nauru. Birth Notification and Certificate and Cause of Death Notiffication. |
| Netherlands | stillbirths | Eurostat Database |
| North Macedonia | stillbirths | Eurostat Database |
| Norway | stillbirths | UNICEF data call. Statisitcs Norway. Medical Birth Registry of Norway and Norwegian Cause of Death Registry. |
| Oman | stillbirths | UNICEF data call. Government of Oman. Vital statistics and civil registration and Health Information System. |
| Panama | stillbirths | UNICEF data call. Panama Ministry of Health. Vital statistics and civil registration. |
| Philippines | stillbirths | UNICEF data call. Philippine Statistics Authority - CRVS. Data submitted to UNICEF via e-mail |
| Poland | stillbirths | Eurostat Database |
| Portugal | stillbirths | UNICEF data call. Statistics Portugal. Civil Registration and Vital Statistics. |
| Qatar | stillbirths | UNICEF data call. Qatar Ministry of Public Health. Births and Deaths Registration. |
| Romania | stillbirths | UNICEF data call. National Institute of Statistics of Romania. Vital registration. https://insse.ro/cms/en |
| Serbia | stillbirths | UNICEF data call. Statistical Office of the Republic of Serbia. Administrative data source. Birth and death register. |
| Singapore | stillbirths | UNICEF data call. Singapore Ministry of Health. Registry of Births and Deaths. |
| Slovakia | stillbirths | UNICEF data call. Statistical Office of the Slovak Republic, Registry offices. Statistical Report on Birth and Medical Report and the Statistical Report on Death. |
| Slovenia | stillbirths | UNICEF data call. National Institute of Public Health - CRVS |

| Country / Territory | Measure | Source |
|---|---|---|
| South Africa | stillbirths | UNICEF data call. Rapid Mortality Surveillance, supplied by R. Dorrington. |
| South Korea | stillbirths | UNICEF data call. Statisitcs Korea. Civil registration system. https://kosis.kr, https://mdis.kostat.go.kr. |
| Spain | stillbirths | Eurostat Database |
| Sri Lanka | stillbirths | UNICEF data call. Perinatal death Surveillance unit -Family Health Bureau - CRVS |
| Sweden | stillbirths | UNICEF data call. Swedish National Board of Health and Welfare. The Swedish Medical Birth Register and the Swedish Cause of Death Register. |
| Switzerland | stillbirths | UNICEF data call. Federal statistical office, Switzerland. Vita Statistics, BEVNAT. https://www.bfs.admin.ch/bfs/en/home/statistics/population/births-deaths/. |
| Turkey | stillbirths | UNICEF data call. Turkish Statistical Institute (TURKSTAT) and Turkish Minsitry of Health. Central Civil Registration System (MERNÃ„Â°S) and Death Notification System (OBS). |
| Uganda | stillbirths | Health Management Information System (HMIS) |
| Ukraine | stillbirths | UNICEF data call. State Statistics Service of Ukraine. Administrative data of vital records. |
| United Kingdom | stillbirths | UNICEF data call. Mothers and Babies: Reducing Risk through Audits and Confidential Enquiries across the UK (MBRRACE-UK) - CRSV |
| Uruguay | stillbirths | UNICEF data call. Uruguay Ministry of Health. Vital Statistics. https://uins.msp.gub.uy/. |
| USA | stillbirths | UNICEF data call. Centers for Disease Control and Prevention of the United States (CDC) - CRVS. Data submitted to UNICEF via e-mail |
| Zambia | stillbirths | Health Management Information System (HMIS) |
| Zimbabwe | stillbirths | Health Management Information System (HMIS) |
| Albania | neonatal | UNPD - Eurostat Database |
| Andorra | neonatal | UNICEF data call. Andorra Ministry of Health. Mortality and birth register of https://www.salut.ad/temes-de-salut/natalitat-i-mortalitat. |
| Argentina | neonatal | UNICEF data call. Argentina Ministerio de Salud - CRVS. Data submitted to UNICEF via e-mail |
| Australia | neonatal | UNICEF data call. Australian Bureau of Statistics. Vital statistics and civil registration (CRVS). https://www. abs.gov.au. |
| Austria | neonatal | Eurostat Database |
| Bangladesh | neonatal | Health Management Information System (HMIS) |
| Belgium | neonatal | Eurostat Database |
| Bosnia and Herzegovina | neonatal | UNICEF data call. Agency for Statistics of Bosnia and Herzegovina. Civil registration and vital statistics. |
| Brazil | neonatal | UNICEF data call. Ministerio da Saude - Datasus https://datasus.saude.gov.br/ |
| Bulgaria | neonatal | UNICEF data call. National Statistical Institute of Bulgaria, Information System Demography. https://www.czso.cz/csu/czso/home. |
| Burkina Faso | neonatal | Health Management Information System (HMIS) |
| Burundi | neonatal | Health Management Information System (HMIS) |
| Canada | neonatal | UNICEF data call. Statistics Canada. Vital statistics and civil registration (CRVS). https://www23.statcan.gc.ca/ |
| Colombia | neonatal | UNICEF data call. National Administrative Department of Statistics (DANE) - CRVS. Data submitted to UNICEF via e-mail |
| Costa Rica | neonatal | UNPD - UN IGME Total Under-5 Mortality Rate, Infant Mortality Rate and Neonatal mortality rate database 2021. |
| Croatia | neonatal | Eurostat Database |
| Cuba | neonatal | UNPD - UN IGME Total Under-5 Mortality Rate, Infant Mortality Rate and Neonatal mortality rate database 2021. |
| Cyprus | neonatal | UNICEF data call. Republic of Cyprus. Ministry of Health. Medical Birth and Death Registry. |
| Czechia | neonatal | UNICEF data call. Czech Statistical Office. CRVS-statistical reports on birth/death. https://www.czso.cz/csu/czso/home. |
| Denmark | neonatal | Eurostat Database |
| Ecuador | neonatal | UNICEF data call. Instituto Nacional de EstadÃƒÂstica y Censos (INEC) - CRVS. Data submitted to UNICEF via e-mail |
| Estonia | neonatal | UNICEF data call. National Institute for Health Development, Estonia. Estonian Medical Birth Registry and Estonian Abortion Registry and Estonian Causes of Death Registry |
| Eswatini | neonatal | Health Management Information System (HMIS) |
| Ethiopia | neonatal | Health Management Information System (HMIS) |
| Finland | neonatal | UNICEF data call. Statistics Finland. Digital and Population Data Services Agency. |
| France | neonatal | Eurostat Database |
| Georgia | neonatal | UNICEF data call. National Statistics Office of Georgia (Geostat). Administrative data. https://www.geostat.ge/en/modules/categories/316/population-and-demography. |
| Germany | neonatal | Eurostat Database |

| Country / Territory | Measure | Source |
|---|---|---|
| Greece | neonatal | UNICEF data call. Hellenic Statistical Authority - CRVS. Data submitted to UNICEF via email. |
| Hungary | neonatal | UNICEF data call. Hungarian Central Statistical Office. Department of Population Statistics. https://www.ksh.hu/. |
| Iceland | neonatal | UNICEF data call. Icelandic Medical Birth Registry |
| India | neonatal | Ministry of Health and Family Welfare of India. Health Management Information System (HMIS). https://hmis.mohfw.gov.in |
| Ireland | neonatal | Eurostat Database |
| Israel | neonatal | UNICEF data call. Israel Central Bureau of Statistics. Population register. www.cbs.gov.il. |
| Italy | neonatal | Eurostat Database |
| Japan | neonatal | UNICEF data call. Ministry of Health of Japan, Labour and Welfare. Vital Statistics of Japan. https://www.e-stat.go.jp/. |
| Kazakhstan | neonatal | UNPD - UN IGME Total Under-5 Mortality Rate, Infant Mortality Rate and Neonatal mortality rate database 2021. |
| Kenya | neonatal | Health Management Information System (HMIS) |
| Kuwait | neonatal | UNICEF data call. Kuwait Central Statistical Bureau and Kuwait Ministry of Health. Vital Statistics. |
| Latvia | neonatal | UNICEF data call. Latvia Ministry of Health, The Centre for Disease Prevention and Control of Latvia. Medical Birth Register and Register of Causes of Death. |
| Liberia | neonatal | Health Management Information System (HMIS) |
| Lithuania | neonatal | UNICEF data call. Statistics Lithuania. Population register. |
| Luxembourg | neonatal | Eurostat Database |
| Madagascar | neonatal | Health Management Information System (HMIS) |
| Malawi | neonatal | Health Management Information System (HMIS) |
| Malaysia | neonatal | UNICEF data call. Ministry of Health of Malaysia. Department of National Registration. |
| Maldives | neonatal | UNICEF data call. Maldives Ministry of Health. Vital registration. |
| Malta | neonatal | UNICEF data call. Ministry for Health and National Statistics Office of Malta. Malta National Birth and Mortality Registry. Data submitted to UNICEF via email. |
| Mauritius | neonatal | UNICEF data call. Statistics Mauritius, Vital statistics and civil registration. |
| Mexico | neonatal | Instituto Nacional de Estadistica y Geografia (INEGI) - CRVS |
| Moldova | neonatal | UNPD - UN IGME Total Under-5 Mortality Rate, Infant Mortality Rate and Neonatal mortality rate database 2021. |
| Mongolia | neonatal | UNICEF data call. Mongolia Ministry of Health. National Center for Health Development. Administrative records. |
| Montenegro | neonatal | UNPD - UN IGME Total Under-5 Mortality Rate, Infant Mortality Rate and Neonatal mortality rate database 2021. |
| Mozambique | neonatal | Countrywide Mortality Surveillance for Action (COMSA) |
| Nauru | neonatal | UNICEF data call. Government of the Republic of Nauru. Birth Notification and Certificate and Cause of Death Notifification. |
| Netherlands | neonatal | Eurostat Database |
| North Macedonia | neonatal | UNPD - Eurostat Database |
| Norway | neonatal | UNICEF data call. Statisitcs Norway. Medical Birth Registry of Norway and Norwegian Cause of Death Registry. |
| Oman | neonatal | UNICEF data call. Government of Oman. Vital statistics and civil registration and Health Information System. |
| Panama | neonatal | UNICEF data call. Panama Ministry of Health. Vital statistics and civil registration. |
| Philippines | neonatal | UNICEF data call. Philippine Statistics Authority - CRVS. Data submitted to UNICEF via e-mail |
| Poland | neonatal | UNICEF data call. Statisitics Poland. Civil Registration and Vital Statistics. https://stat.gov.pl/. |
| Portugal | neonatal | UNICEF data call. Statistics Portugal. Civil Registration and Vital Statistics. |
| Qatar | neonatal | UNICEF data call. Qatar Ministry of Public Health. Births and Deaths Registration. |
| Romania | neonatal | UNICEF data call. National Institute of Statistics of Romania. Vital registration. https://insse.ro/cms/en |
| Russia | neonatal | UNICEF data call. Russian Federal State Statistics Service. Vital registration data. Data submitted to UNICEF via email. |
| Serbia | neonatal | UNICEF data call. Statistical Office of the Republic of Serbia. Administrative data source. Birth and death register. |
| Singapore | neonatal | UNICEF data call. Singapore Ministry of Health. Registry of Births and Deaths. |
| Slovakia | neonatal | UNICEF data call. Statistical Office of the Slovak Republic, Registry offices. Statistical Report on Birth and Medical Report and the Statistical Report on Death. |
| Slovenia | neonatal | UNICEF data call. National Institute of Public Health - CRVS |
| South Africa | neonatal | UNICEF data call. Rapid Mortality Surveillance, supplied by R. Dorrington. |

| Country / Territory | Measure | Source |
|---|---|---|
| South Korea | neonatal | UNICEF data call. Statisitcs Korea. Civil registration system. https://kosis.kr, https://mdis.kostat.go.kr. |
| Spain | neonatal | Eurostat Database |
| Sri Lanka | neonatal | UNICEF data call. Perinatal death Surveillance unit -Family Health Bureau - CRVS |
| Sweden | neonatal | UNICEF data call. Swedish National Board of Health and Welfare. The Swedish Medical Birth Register and the Swedish Cause of Death Register, |
| Switzerland | neonatal | UNICEF data call. Federal statistical office, Switzerland. Vita Statistics, BEVNAT. |
| Turkey | neonatal | UNICEF data call. Turkish Statistical Institute (TURKSTAT) and Turkish Minsitry of Health. Central Civil Registration System (MERNÃ„Â°S) and Death Notification System (OBS). |
| Uganda | neonatal | Health Management Information System (HMIS) |
| Ukraine | neonatal | UNICEF data call. State Statistics Service of Ukraine. Administrative data of vital records. |
| United Kingdom | neonatal | UNICEF data call. Mothers and Babies: Reducing Risk through Audits and Confidential Enquiries across the UK (MBRRACE-UK) - CRSV |
| Uruguay | neonatal | UNICEF data call. Uruguay Ministry of Health. Vital Statistics. https://uins.msp.gub.uy/. |
| USA | neonatal | UNICEF data call. Centers for Disease Control and Prevention of the United States (CDC) - CRVS. Data submitted to UNICEF via e-mail |
| Uzbekistan | neonatal | UNICEF data call. Statistics Agency under the President of the Republic of Uzbekistan. Department of Demography and Labor Statistics. Vital registration. |
| Zambia | neonatal | Health Management Information System (HMIS) |
| Zimbabwe | neonatal | Health Management Information System (HMIS) |
| Albania | infant | Eurostat Database |
| American Samoa | infant | UNPD - Demographic Yearbook |
| Andorra | infant | UNPD - WHO All-Cause Mortality Data Call |
| Antigua and Barbuda | infant | WHO Mortality Database |
| Argentina | infant | UNPD - WHO Mortality Data base |
| Aruba | infant | UNPD - Demographic Yearbook |
| Australia | infant | UNICEF data call. Australian Bureau of Statistics. Vital statistics and civil registration (CRVS). https://www. abs.gov.au. |
| Austria | infant | WHO Mortality Database |
| Bangladesh | infant | Bangladesh Bureau of Statistics - CRVS |
| Belgium | infant | Human Mortality Database (HMD) |
| Belize | infant | UNPD - Demographic Yearbook |
| Bermuda | infant | UNPD - Demographic Yearbook |
| Bosnia and Herzegovina | infant | UNICEF data call. Agency for Statistics of Bosnia and Herzegovina. Civil registration and vital statistics. |
| Brazil | infant | Ministerio da Saude - Datasus https://datasus.saude.gov.br/ |
| Brunei Darussalam | infant | UNPD - Demographic Yearbook |
| Bulgaria | infant | UNPD - WHO All-Cause Mortality Data Call |
| Canada | infant | UNPD - WHO All-Cause Mortality Data Call |
| Chile | infant | UNPD - WHO All-Cause Mortality Data Call |
| China | infant | National Bureau of Statistics of China - SVR |
| Colombia | infant | UNICEF data call. National Administrative Department of Statistics (DANE) - CRVS. Data submitted to UNICEF via e-mail |
| Costa Rica | infant | UNPD - WHO All-Cause Mortality Data Call |
| Croatia | infant | UNPD - WHO All-Cause Mortality Data Call |
| Cuba | infant | WHO Mortality Database |
| Cyprus | infant | UNICEF data call. Republic of Cyprus. Ministry of Health. Medical Birth and Death Registry. |
| Czechia | infant | Human Mortality Database (HMD) |
| Denmark | infant | Human Mortality Database (HMD) |
| Dominica | infant | UNPD - WHO Mortality Data base |
| Dominican Republic | infant | UNPD - Demographic Yearbook |
| Ecuador | infant | WHO Mortality Database |
| Egypt | infant | UNPD - Demographic Yearbook |
| Estonia | infant | WHO Mortality Database |
| Finland | infant | Human Mortality Database (HMD) |
| France | infant | Eurostat Database |
| French Guiana | infant | UNPD - Demographic Yearbook |
| French Polynesia | infant | UNPD - Demographic Yearbook |
| Georgia | infant | UNICEF data call. National Statistics Office of Georgia (Geostat). Administrative data. https://www.geostat.ge/en/modules/categories/316/population-and-demography. |
| Germany | infant | Eurostat Database |

| Country / Territory | Measure | Source |
|---|---|---|
| Greece | infant | WHO Mortality Database |
| Grenada | infant | WHO Mortality Database |
| Guatemala | infant | WHO Mortality Database |
| Honduras | infant | UNICEF data call. Secretaria de Salud of Honduras. Area Estadistica de la Salud. Data submitted to UNICEF via e-mail |
| Hong Kong - China SAR | infant | UNPD - Demographic Yearbook |
| Hungary | infant | UNICEF data call. Hungarian Central Statistical Office. Department of Population Statistics. https://www.ksh.hu/. |
| Iceland | infant | UNPD - Demographic Yearbook |
| India | infant | Office of the Registrar General & Census Commissioner, India (ORGI) - Sample Registration System (SRS) |
| Iran | infant | UNPD - Demographic Yearbook |
| Ireland | infant | Human Mortality Database (HMD) |
| Israel | infant | UNICEF data call. Israel Central Bureau of Statistics. Population register. www.cbs.gov.il. |
| Italy | infant | Short-term mortality fluctuations database (STMF) |
| Japan | infant | Human Mortality Database (HMD) |
| Kazakhstan | infant | UNPD - Demographic Yearbook |
| Kuwait | infant | UNICEF data call. Kuwait Central Statistical Bureau and Kuwait Ministry of Health. Vital Statistics. |
| Kyrgyzstan | infant | UNPD - Demographic Yearbook |
| Latvia | infant | UNICEF data call. Latvia Ministry of Health, The Centre for Disease Prevention and Control of Latvia. Medical Birth Register and Register of Causes of Death. https://statistika.spkc.gov.lv/. |
| Lebanon | infant | WHO Mortality Database |
| Liechtenstein | infant | Eurostat Database |
| Lithuania | infant | UNICEF data call. Statistics Lithuania. Population register. |
| Luxembourg | infant | Human Mortality Database (HMD) |
| Malaysia | infant | UNICEF data call. Ministry of Health of Malaysia. Department of National Registration. |
| Maldives | infant | UNICEF data call. Maldives Ministry of Health. Vital registration. |
| Malta | infant | Eurostat Database |
| Martinique | infant | UNPD - Demographic Yearbook |
| Mauritius | infant | UNICEF data call. Statistics Mauritius, Vital statistics and civil registration. |
| Mayotte | infant | UNPD - Demographic Yearbook |
| Mexico | infant | UNPD - WHO Mortality Data base |
| Moldova | infant | UNPD - WHO All-Cause Mortality Data Call |
| Mongolia | infant | UNICEF data call. Mongolia Ministry of Health. National Center for Health Development. Administrative records. |
| Montenegro | infant | Eurostat Database |
| Montserrat | infant | WHO Mortality Database |
| Nauru | infant | UNICEF data call. Government of the Republic of Nauru. Birth Notification and Certificate and Cause of Death Notifification. |
| Netherlands | infant | WHO Mortality Database |
| New Zealand | infant | Human Mortality Database (HMD) |
| Nicaragua | infant | UNPD - WHO Mortality Data base |
| North Macedonia | infant | UNPD - WHO Mortality Data base |
| Norway | infant | Human Mortality Database (HMD) |
| Oman | infant | UNPD - Demographic Yearbook |
| Panama | infant | UNICEF data call. Panama Ministry of Health. Vital statistics and civil registration. |
| Paraguay | infant | UNPD - WHO Mortality Data base |
| Peru | infant | WHO Mortality Database |
| Philippines | infant | UNICEF data call. Philippine Statistics Authority - CRVS. Data submitted to UNICEF via e-mail |
| Poland | infant | UNPD - WHO All-Cause Mortality Data Call |
| Portugal | infant | UNICEF data call. Statistics Portugal. Civil Registration and Vital Statistics. |
| Puerto Rico | infant | UNPD - Demographic Yearbook |
| Qatar | infant | UNPD - Demographic Yearbook |
| Reunion | infant | UNPD - Demographic Yearbook |
| Romania | infant | UNPD - WHO All-Cause Mortality Data Call |
| Russia | infant | UNICEF data call. Russian Federal State Statistics Service. Vital registration data. Data submitted to UNICEF via email. |
| Saint Lucia | infant | UNPD - WHO Mortality Data base |
| Serbia | infant | WHO Mortality Database |
| Seychelles | infant | UNPD - Demographic Yearbook |

| Country / Territory | Measure | Source |
|---|---|---|
| Singapore | infant | UNPD - Demographic Yearbook |
| Slovakia | infant | UNICEF data call. Statistical Office of the Slovak Republic, Registry offices. Statistical Report on Birth and Medical Report and the Statistical Report on Death. |
| Slovenia | infant | WHO Mortality Database |
| South Africa | infant | UNICEF data call. Rapid Mortality Surveillance, supplied by R. Dorrington. |
| South Korea | infant | UNICEF data call. Statisitcs Korea. Civil registration system. https://kosis.kr, https://mdis.kostat.go.kr. |
| Spain | infant | WHO Mortality Database |
| Sri Lanka | infant | UNICEF data call. Perinatal death Surveillance unit -Family Health Bureau - CRVS |
| State of Palestine | infant | UNPD - Demographic Yearbook |
| Suriname | infant | UNPD - Demographic Yearbook |
| Sweden | infant | Human Mortality Database (HMD) |
| Switzerland | infant | UNICEF data call. Federal statistical office, Switzerland. Vita Statistics, BEVNAT. https://www.bfs.admin.ch/bfs/en/home/statistics/population/births-deaths/. |
| Thailand | infant | UNICEF data call. Thailand Ministry of Interior. Civil registration and vital statistics (CRVS). https://stat.bora.dopa.go.th/stat/statnew/statMenu/newStat/stat/. |
| Turkey | infant | UNICEF data call. Turkish Statistical Institute (TURKSTAT) and Turkish Minsitry of Health. Central Civil Registration System (MERNÃ„Â°S) and Death Notification System (OBS). |
| Turks and Caicos Islands | infant | UNPD - Demographic Yearbook |
| Ukraine | infant | UNICEF data call. State Statistics Service of Ukraine. Administrative data of vital records. |
| United Arab Emirates | infant | UNPD - Demographic Yearbook |
| United Kingdom | infant | Human Mortality Database (HMD) |
| Uruguay | infant | UNICEF data call. Uruguay Ministry of Health. Vital Statistics. https://uins.msp.gub.uy/. |
| USA | infant | Human Mortality Database (HMD) |
| Uzbekistan | infant | UNPD - Demographic Yearbook |
| Afghanistan | deaths 0-24 | Health Management Information System (HMIS) |
| Albania | deaths 0-24 | Eurostat Database |
| American Samoa | deaths 0-24 | UNPD - Demographic Yearbook |
| Andorra | deaths 0-24 | UNPD - WHO All-Cause Mortality Data Call |
| Antigua and Barbuda | deaths 0-24 | WHO Mortality Database |
| Argentina | deaths 0-24 | UNPD - WHO Mortality Data base |
| Aruba | deaths 0-24 | UNPD - Demographic Yearbook |
| Australia | deaths 0-24 | WHO Mortality Database |
| Austria | deaths 0-24 | WHO Mortality Database |
| Bangladesh | deaths 0-24 | Bangladesh Bureau of Statistics - Sample Vital Statistics. |
| Belgium | deaths 0-24 | Human Mortality Database (HMD) |
| Belize | deaths 0-24 | UNPD - Demographic Yearbook |
| Bermuda | deaths 0-24 | UNPD - Demographic Yearbook |
| Bosnia and Herzegovina | deaths 0-24 | UNICEF data call. Agency for Statistics of Bosnia and Herzegovina. Civil registration and vital statistics. |
| Brazil | deaths 0-24 | Ministerio da Saude - Datasus https://datasus.saude.gov.br/ |
| Brunei Darussalam | deaths 0-24 | UNPD - Demographic Yearbook |
| Bulgaria | deaths 0-24 | UNPD - WHO All-Cause Mortality Data Call |
| Burkina Faso | deaths 0-24 | Health Management Information System (HMIS) |
| Canada | deaths 0-24 | UNPD - WHO All-Cause Mortality Data Call |
| Chile | deaths 0-24 | UNPD - WHO All-Cause Mortality Data Call |
| China | deaths 0-24 | National Bureau of Statistics of China - SVR |
| Colombia | deaths 0-24 | UNICEF data call. National Administrative Department of Statistics of Colombia (DANE) - CRVS. |
| Costa Rica | deaths 0-24 | UNPD - WHO All-Cause Mortality Data Call |
| Croatia | deaths 0-24 | Short-term mortality fluctuations database (STMF) |
| Cuba | deaths 0-24 | WHO Mortality Database |
| Cyprus | deaths 0-24 | Eurostat Database |
| Czechia | deaths 0-24 | Short-term mortality fluctuations database (STMF) |
| Denmark | deaths 0-24 | Human Mortality Database (HMD) |
| Dominica | deaths 0-24 | UNPD - WHO Mortality Data base |
| Dominican Republic | deaths 0-24 | UNPD - Demographic Yearbook |
| Ecuador | deaths 0-24 | WHO Mortality Database |
| Egypt | deaths 0-24 | UNPD - Demographic Yearbook |
| Estonia | deaths 0-24 | Short-term mortality fluctuations database (STMF) |
| Ethiopia | deaths 0-24 | Health Management Information System (HMIS) |

| Country / Territory | Measure | Source |
|---|---|---|
| Finland | deaths 0-24 | Short-term mortality fluctuations database (STMF) |
| France | deaths 0-24 | Eurostat Database |
| French Guiana | deaths 0-24 | UNPD - Demographic Yearbook |
| French Polynesia | deaths 0-24 | UNPD - Demographic Yearbook |
| Georgia | deaths 0-24 | UNICEF data call. National Statistics Office of Georgia (Geostat). Administrative data. |
| Germany | deaths 0-24 | UNPD - WHO Mortality Data base |
| Greece | deaths 0-24 | Short-term mortality fluctuations database (STMF) |
| Grenada | deaths 0-24 | WHO Mortality Database |
| Guatemala | deaths 0-24 | WHO Mortality Database |
| Honduras | deaths 0-24 | UNICEF data call. Secretaria de Salud of Honduras. Area Estadistica de la Salud. Data submitted to UNICEF via e-mail |
| Hong Kong - China SAR | deaths 0-24 | UNPD - Demographic Yearbook |
| Hungary | deaths 0-24 | Eurostat Database |
| Iceland | deaths 0-24 | Eurostat Database |
| India | deaths 0-24 | Ministry of Health and Family Welfare of India. Health Management Information System (HMIS). https://hmis.mohfw.gov.in |
| Iran | deaths 0-24 | UNPD - Demographic Yearbook |
| Ireland | deaths 0-24 | Human Mortality Database (HMD) |
| Israel | deaths 0-24 | UNICEF data call. Israel Central Bureau of Statistics. Population register. www.cbs.gov.il. |
| Italy | deaths 0-24 | Short-term mortality fluctuations database (STMF) |
| Japan | deaths 0-24 | Human Mortality Database (HMD) |
| Kazakhstan | deaths 0-24 | UNPD - Demographic Yearbook |
| Kenya | deaths 0-24 | Health Management Information System (HMIS) |
| Kuwait | deaths 0-24 | UNICEF data call. Kuwait Central Statistical Bureau and Kuwait Ministry of Health. Vital Statistics. |
| Kyrgyzstan | deaths 0-24 | UNPD - Demographic Yearbook |
| Latvia | deaths 0-24 | Eurostat Database |
| Lebanon | deaths 0-24 | WHO Mortality Database |
| Liberia | deaths 0-24 | Health Management Information System (HMIS) |
| Libya | deaths 0-24 | UNICEF data call. Libyan Civil Registration Authority. Electronic system based on actual deaths and births registered by date. |
| Liechtenstein | deaths 0-24 | Eurostat Database |
| Lithuania | deaths 0-24 | Short-term mortality fluctuations database (STMF) |
| Luxembourg | deaths 0-24 | Short-term mortality fluctuations database (STMF) |
| Malawi | deaths 0-24 | Health Management Information System (HMIS) |
| Malaysia | deaths 0-24 | UNICEF data call. Ministry of Health of Malaysia. Department of National Registration. |
| Maldives | deaths 0-24 | UNICEF data call. Maldives Ministry of Health. Vital registration. |
| Malta | deaths 0-24 | Eurostat Database |
| Martinique | deaths 0-24 | UNPD - Demographic Yearbook |
| Mauritius | deaths 0-24 | UNICEF data call. Statistics Mauritius, Vital statistics and civil registration. |
| Mayotte | deaths 0-24 | UNPD - Demographic Yearbook |
| Mexico | deaths 0-24 | UNPD - WHO Mortality Data base |
| Moldova | deaths 0-24 | UNPD - WHO All-Cause Mortality Data Call |
| Mongolia | deaths 0-24 | UNICEF data call. Mongolia Ministry of Health. National Center for Health Development. Administrative records. |
| Montenegro | deaths 0-24 | Eurostat Database |
| Montserrat | deaths 0-24 | WHO Mortality Database |
| Mozambique | deaths 0-24 | Countrywide Mortality Surveillance for Action (COMSA) |
| Nauru | deaths 0-24 | UNICEF data call. Government of the Republic of Nauru. Birth Notification and Certificate and Cause of Death Notiffication. |
| Netherlands | deaths 0-24 | Short-term mortality fluctuations database (STMF) |
| New Zealand | deaths 0-24 | UNPD - Demographic Yearbook |
| Nicaragua | deaths 0-24 | UNPD - WHO Mortality Data base |
| North Macedonia | deaths 0-24 | UNPD - WHO Mortality Data base |
| Norway | deaths 0-24 | Human Mortality Database (HMD) |
| Oman | deaths 0-24 | UNPD - Demographic Yearbook |
| Panama | deaths 0-24 | UNICEF data call. Panama Ministry of Health. Vital statistics and civil registration. |
| Paraguay | deaths 0-24 | UNPD - WHO All-Cause Mortality Data Call |
| Peru | deaths 0-24 | WHO Mortality Database |
| Philippines | deaths 0-24 | UNICEF data call. Philippine Statistics Authority - CRVS. Data submitted to UNICEF via e-mail |
| Poland | deaths 0-24 | Short-term mortality fluctuations database (STMF) |
| Portugal | deaths 0-24 | Short-term mortality fluctuations database (STMF) |

| Country / Territory | Measure | Source |
|---|---|---|
| Puerto Rico | deaths 0-24 | UNPD - Demographic Yearbook |
| Qatar | deaths 0-24 | UNPD - Demographic Yearbook |
| Reunion | deaths 0-24 | UNPD - Demographic Yearbook |
| Romania | deaths 0-24 | Eurostat Database |
| Russia | deaths 0-24 | Short-term mortality fluctuations database (STMF) |
| Saint Lucia | deaths 0-24 | UNPD - WHO Mortality Data base |
| Serbia | deaths 0-24 | WHO Mortality Database |
| Seychelles | deaths 0-24 | UNPD - Demographic Yearbook |
| Singapore | deaths 0-24 | UNPD - Demographic Yearbook |
| Slovakia | deaths 0-24 | Eurostat Database |
| Slovenia | deaths 0-24 | Short-term mortality fluctuations database (STMF) |
| South Africa | deaths 0-24 | UNICEF data call. Rapid Mortality Surveillance, supplied by R. Dorrington. |
| South Korea | deaths 0-24 | Human Mortality Database (HMD) |
| Spain | deaths 0-24 | WHO Mortality Database |
| Sri Lanka | deaths 0-24 | UNICEF data call. Perinatal death Surveillance unit -Family Health Bureau - CRVS |
| State of Palestine | deaths 0-24 | UNPD - Demographic Yearbook |
| Suriname | deaths 0-24 | UNPD - Demographic Yearbook |
| Sweden | deaths 0-24 | Human Mortality Database (HMD) |
| Switzerland | deaths 0-24 | UNICEF data call. Federal statistical office, Switzerland. Vita Statistics, BEVNAT. |
| Taiwan - Province of China | deaths 0-24 | Short-term mortality fluctuations database (STMF) |
| Thailand | deaths 0-24 | UNICEF data call. Thailand Ministry of Interior. Civil registration and vital statistics (CRVS). |
| Turkey | deaths 0-24 | UNICEF data call. Turkish Statistical Institute (TURKSTAT) and Turkish Minsitry of Health. Central Civil Registration System (MERNÃ„Â°S) and Death Notification System (OBS). |
| Turks and Caicos Islands | deaths 0-24 | UNPD - Demographic Yearbook |
| Uganda | deaths 0-24 | Health Management Information System (HMIS) |
| Ukraine | deaths 0-24 | UNICEF data call. State Statistics Service of Ukraine. Administrative data of vital records. |
| United Arab Emirates | deaths 0-24 | UNPD - Demographic Yearbook |
| United Kingdom | deaths 0-24 | Human Mortality Database (HMD) |
| Uruguay | deaths 0-24 | UNICEF data call. Uruguay Ministry of Health. Vital Statistics. https://uins.msp.gub.uy/. |
| USA | deaths 0-24 | Human Mortality Database (HMD) |
| Uzbekistan | deaths 0-24 | UNPD - Demographic Yearbook |
